# Heartbeat-evoked neural response abnormalities in generalized anxiety disorder during peripheral adrenergic stimulation

**DOI:** 10.1101/2023.06.09.23291166

**Authors:** Charles Verdonk, Adam R. Teed, Evan J. White, Xi Ren, Jennifer L. Stewart, Martin P. Paulus, Sahib S. Khalsa

## Abstract

Hyperarousal symptoms in generalized anxiety disorder (GAD) are often incongruent with the observed physiological state, suggesting that abnormal processing of interoceptive signals is a characteristic feature of the disorder. To examine the neural mechanisms underlying interoceptive dysfunction in GAD, we evaluated whether adrenergic modulation of cardiovascular signaling differentially affects the heartbeat evoked potential (HEP), an electrophysiological marker of cardiac interoception, during concurrent electroencephalogram and functional magnetic resonance imaging (EEG-fMRI) scanning. Intravenous infusions of the peripheral adrenergic agonist isoproterenol (0.5 and 2.0 micrograms, μg) were administered in a randomized, double-blinded and placebo-controlled fashion to dynamically perturb the cardiovascular system while recording the associated EEG-fMRI responses. During the 0.5 μg isoproterenol infusion, the GAD group (n=24) exhibited significantly larger changes in HEP amplitude in an opposite direction than the HC group (n=24). In addition, the GAD group showed significantly larger absolute HEP amplitudes than HC during saline infusions, when cardiovascular tone did not increase. No significant group differences in HEP amplitude were identified during the 2.0 μg isoproterenol infusion. Using analyzable blood oxygenation level dependent fMRI data from participants with concurrent EEG-fMRI data (21 GAD and 21 HC), we found that the aforementioned HEP effects were uncorrelated with fMRI signals in the insula, ventromedial prefrontal cortex, dorsal anterior cingulate cortex, amygdala, and somatosensory cortex, brain regions implicated in cardiac signal processing according to prior fMRI studies. These findings provide additional evidence of dysfunctional cardiac interoception in GAD and identify neural processes at the electrophysiological level that may be independent from blood oxygen level–dependent responses during peripheral adrenergic stimulation.

## INTRODUCTION

Generalized anxiety disorder (GAD) is a prevalent, chronic, and debilitating psychiatric condition characterized by excessive and uncontrollable worry about everyday life events [1, 2]. Elevated sympathetic arousal symptoms such as heart palpitations and shortness of breath are commonly reported by individuals with GAD [3], although diagnostic formulation in the DSM preferentially emphasizes other bodily symptoms including fatigue, headache, and muscle tension. The pathophysiology of GAD remains poorly understood, but it has been suggested that abnormal physiological arousal plays a crucial role in the development and maintenance of the disorder [4, 5]. The affective and sympathetic arousal symptoms reported by individuals with GAD are often mismatched with their actual physiological state [6, 7], consistent with the idea that dysfunctional interoception is a characteristic feature of the disorder [8, 9]. In this context, identifying the interoceptive mechanisms underlying GAD could provide insight into the pathophysiology of the disorder and open up new avenues for treatment.

Functional neuroimaging studies have revealed a network of brain regions, including the insular, somatosensory, and ventromedial prefrontal cortices (vmPFC), that plays a central role in the integration and representation of resting cardiovascular signals [10, 11]. Some of these brain regions, specifically the insula and vmPFC, have been associated with awareness of sympathetically modulated cardiac sensations such as those induced by the administration of isoproterenol, a peripheral beta-adrenergic agonist akin to adrenaline [12-14]. In line with these findings, we recently observed abnormal physiological, neural and subjective responses to adrenergic stimulation in women with GAD relative to healthy comparisons (HC), using a within-subjects crossover design with isoproterenol (0.5 and 2.0 micrograms, μg) and saline during concurrent electroencephalogram and functional MRI (EEG-fMRI) scanning [15]. Our fMRI results demonstrated that the participants with GAD exhibited vmPFC hypoactivation relative to HCs, in combination with heightened ratings of cardiorespiratory sensation and anxiety. These findings suggested that a disrupted central regulatory threshold in response to cardiac signals in women with GAD may be mediated by the vmPFC, implying a top-down disruption of physiological state that promotes anxious responses [15]. However, we also observed heightened heart rate (HR) responses during isoproterenol stimulation in the GAD group, raising the possibility of increased peripheral adrenergic sensitivity as an additional contributor to interoceptive dysfunction in the disorder.

The neurophysiological basis of cardiac interoception can be investigated by probing heartbeat-related neural responses, specifically via the heartbeat-evoked potential (HEP), an evoked response potential measured via EEG [16]. The HEP has been proposed as an electrophysiological biomarker of cardiac interoceptive processing based on meta-analytic evidence of associations between its amplitude and behavioral measures of cardiac interoception (i.e., performance on heartbeat perception tasks), experimentally-induced changes in cardiovascular arousal using emotionally-relevant stimuli (e.g., angry face presentation), and manipulation of attention to heartbeat signals [17]. Only one study has previously investigated the HEP and modulation of its amplitude in GAD. In that study, Pang et al. [18] evaluated HEP amplitudes in 25 individuals with GAD and 15 HCs under two physiological resting conditions, eyes-closed and eyes-open, while participants were instructed not to think about anything in particular. The GAD group did not show a significant change in HEP amplitudes during the eyes-closed versus eyes-open conditions, in contrast to the HCs, who exhibited significantly higher HEP amplitudes by comparison. However, anxiety levels in the GAD group were negatively correlated with HEP amplitudes, leading the authors to speculate that the HEP signal reflected an objective marker of anxiety as well as a form of “deficient interoceptive adaptation” [18]. However, probing the HEP while manipulating peripheral cardiovascular arousal is a necessary prerequisite for the evaluation of adaptive bottom-up (i.e., from the heart to the brain) and top-down (i.e., from the brain to the heart) mechanisms of interoceptive signal processing [8, 19]. The protocol of isoproterenol infusion transiently increases HR due to an agonist effect on peripheral beta-adrenergic receptors [20], reflecting an ascending afferent or bottom-up modulation of cardiovascular signals. By contrast, during the saline condition, participants experience the anticipation of infusion-elicited changes in their cardiac sensations that do not occur. In other words, this protocol facilitates a parametric experimental manipulation of both bottom-up (afferent physiological) and top-down (attentional/cognitive) inputs to the heart-brain communication axis.

The present study aimed to investigate the electrophysiological involvement of aberrant bottom-up and top-down processing of cardiovascular signals in GAD using a multilevel approach incorporating the assessment of the HEP alongside simultaneous measurements of subjective and physiological data. The current analysis was informed by our previous observation of vmPFC hypoactivation associated with abnormal cardiac interoception in GAD during low levels of peripheral adrenergic stimulation [15], and applied to the same EEG-fMRI dataset. We hypothesized that, relative to a matched HC group, the GAD group would exhibit greater changes in HEP amplitude during the 0.5 μg dose of isoproterenol versus saline. Indeed, our previous findings have shown hyperarousal in the GAD group during the 0.5 μg dose of isoproterenol [15], and previous HEP studies have demonstrated that the HEP amplitude increases in high arousal conditions [21, 22]. Furthermore, within the predictive coding framework, the HEP amplitude has been suggested to represent interoceptive prediction error, which arises from discrepancies between anticipated (i.e., top-down) and actual (i.e., bottom-up) cardiac inputs [23-26]. Consequently, we expected larger HEP amplitudes in the GAD group relative to the HC group during the saline condition. This would reflect abnormally heightened expectations of infusion-induced cardiovascular changes that did not materialize (i.e., over-weighted expectations driven by top-down mechanisms underlying interoceptive dysfunction [27, 28]). Finally, in a subsample of participants with usable concurrent EEG-fMRI data, we explored associations between HEP signals and blood oxygenation level dependent fMRI signals in brain regions previously observed to show isoproterenol dose-specific fMRI activity changes such as the insula [10] and vmPFC [15], as well as other potential neural sources of HEP activity including the dorsal anterior cingulate cortex (dACC), amygdala, and somatosensory cortex [16], irrespective of diagnosis. Our hypothesis posited that fMRI signals from these regions of interest would be associated with HEP signals across all participants if these were reflecting the same mechanism of interoceptive dysfunction.

## MATERIALS AND METHODS

### Participants

Diagnostic grouping of participants in this crossover randomized clinical trial was based on DSM-IV TR or DSM-5 criteria using the Mini-International Neuropsychiatric Interview [29]. Additional inclusion criteria required patients with GAD to have a score greater than 7 on the Overall Anxiety Severity and Impairment Scale [30] or greater than 10 on the GAD-7 scale [31], indicative of clinically significant anxiety levels. Although comorbid depression and anxiety disorders were allowed, panic disorder was exclusionary to reduce potential dropout associated with isoproterenol-induced panic anxiety [32] (Supplementary Methods 1; Figure S1; Table S1). Selected psychotropic agents were allowed provided there was no change in dosage four weeks before the study. The study was approved by the Western Institutional Review Board. All participants provided written informed consent before participation and were financially compensated for their involvement. ClinicalTrials.gov identifier: #NCT02615119.

### Collection of neurophysiological, cardiovascular, and subjective data

Multilevel data were collected during a multimodal fMRI scanning session (Figure S2A). EEG data were simultaneously collected with BOLD fMRI signals using a 32-channel MR-compatible EEG system (Brain Products GmBH, Germany), whose cap consisted of 32 channels arranged according to the international 10-20 system. One of the 32 channels was devoted to electrocardiogram (ECG) recording via electrode placement on the upper back. EEG and ECG data were acquired at a sampling rate of 5 kHz and a resolution of 0.1 microvolts (μV). In addition to the ECG recording, the cardiac signal was recorded using a MR-compatible photoplethysmogram (PPG) placed on a non-dominant finger and sampled at 40Hz. Respiratory signals were recorded via a thoracic belt. Subjective data were collected via continuously rated changes in the perceived intensity of cardiorespiratory sensations by asking participants to rotate an MRI-compatible dial (Current Designs Inc.) with their dominant hand using a visual analog scale ranging from 0 (“none or normal”) to 10 (“most ever”) (Figure S2B). Demographic matching variables included self-reported gender, age, and measured body mass index (BMI; calculated as weight in kilograms divided by height in meters squared).

### Experimental protocol

The protocol included three experimental conditions defined by the delivery of double-blinded IV bolus infusions of isoproterenol hydrochloride (0.5 and 2 μg) or saline, administered by a nurse located in the scanner room 60 seconds (sec) after the onset of each infusion scan period (Figure S2C). Each infusion condition was repeated once for a total of 6 infusion recordings administered via a randomized crossover procedure (Supplementary Methods 2). On the basis of previous studies implementing bolus isoproterenol infusions [12, 14, 15] we defined four discrete windows of interest as follows (Figure S2C): (1) the anticipatory period (60-80 sec), which began with infusion delivery and preceded the onset of isoproterenol effects, (2) the peak period (80-120 sec) corresponding to the onset and maximal effect, (3) the early recovery period (120-180 sec) corresponding to the initial resolution, and (4) the late recovery period (180-240 sec). The peak and early recovery periods were the primary epochs of interest as these are the windows in which the transient isoproterenol-induced changes in cardiorespiratory state typically occur [12, 14, 15].

### Computation of the Heartbeat Evoked Potential (HEP)

EEG and ECG data were processed using custom scripts in Matlab 2020b (Mathworks^®^) and the EEGLAB toolbox (version 19.0; [33]) for artifact correction, downsampling, bandpass filtering, and re-referencing. The fMRI-related gradient artifact was removed from the EEG and ECG signals using the fMRIb plug-in (version 2.00) for EEGLAB, specifically the command *pop_fmrib_fastr* that implements the Optimal Basis Set (OBS) method (Figure S3) [34]. After downsampling to 250 Hz, the data were band-pass filtered between 0.3 to 30 Hz using the EEGLAB function *eegfilt()*. The cardiac-related ballistocardiogram (BCG) artifact was removed using the OBS method [34], specifically the command *pop_fmrib_pas* of the fMRIb plug-in for EEGLAB (Figure S4). Afterwards, we selected the common average as the reference [35]. Data were epoched with the cardiac R-wave event as the temporal reference, from -100 ms to 650 ms after R-wave onset (Supplementary Methods 3). On average, there were 472 epochs available for analysis per participant per condition (Supplementary Results 1). We subsequently subtracted the mean of the first 50 msec of the epoch (i.e., from -100 to -50 ms before cardiac R-wave event) from the entire signal for each individual average to account for signal drifts commonly characterizing EEG recordings [17, 36]. Then, we implemented an additional step for correcting potential BCG residuals using Independent Component Analysis (Supplementary Methods 3). Independent Component Analysis was also used to correct for eye movement artifacts (Supplementary Methods 3). Finally, based on a previous study showing that repeated measures of the resting-state HEP signal can vary at the individual level [37], we averaged HEP signals from the baseline period before each infusion (first 60 seconds of the recording; Figure S2C) and then subtracted this from period-averaged HEP signals to compensate for within-individual differences between conditions in the baseline interval [38].

### MRI Data Acquisition

T1-weighted anatomical magnetization-prepared, rapid gradient-echo sequence images and T2*-weighted BOLD contrast images via an echoplanar sequence were collected in a 3 Tesla MRI scanner with an 8-channel head coil (GE MR750) as described in [15] (Supplemental Methods 4). Preprocessing and analysis were performed in AFNI [39] using RETROICOR physiological noise-corrected images [40] implemented via custom Matlab code. Individual-level percentage of signal change maps of BOLD response to cardiorespiratory stimulation were generated from residual images by contrasting epoch-averaged signals for isoproterenol epochs (e.g., peak period) against the baseline period. Whole-brain voxelwise t tests using *3dttest++* in AFNI contrasted group-averaged PSC maps for each epoch using a voxelwise threshold of *P*<.001 and a 95% false-positive rate cluster correction.

### Statistical analysis

We used non-parametric cluster permutation to determine if changes in cardiovascular responses induced by isoproterenol had an impact on HEP amplitude as a function of diagnostic group (i.e., GAD vs HC). This data driven statistical technique enabled comparisons between groups across all electrodes at each time point, providing a straightforward way to address the multiple comparison problem (Supplementary Methods 5) [41, 42]. We analyzed group differences (GAD versus HC) within each condition (isoproterenol 0.5 μg, isoproterenol 2 μg, and saline) separately, and between conditions by computing the change in HEP amplitude from the saline to the corresponding isoproterenol infusion (0.5μg or 2μg). Cluster permutation was performed in the time range from 100 to 600 ms after the cardiac R-wave event to avoid contamination by potential residuals of the cardiac field artifact (CFA) and P waves from the ensuing heartbeat in periods of high heart rates. We implemented this statistical method using custom Matlab 2020b scripts and the Fieldtrip toolbox (version 20171022; [43]). Cluster permutation was also used to evaluate isoproterenol’s effects on the time course of heart rate and self-reported cardiorespiratory intensity.

We then explored the relationship between neural, cardiac, and subjective outcomes to examine multilevel associations of interoceptive processing across diagnostic groups. To do this, we computed the average HEP amplitude (avHEP) as the average HEP signal across all electrodes and time points that were included in the HEP cluster identified from the corresponding period of interest (e.g., peak period). The average heart rate (avHR) and the average self-reported cardiorespiratory intensity (avCI) were computed for each period of interest. After examination of data, one HC participant was excluded from fMRI-EEG correlation analysis due to fMRI signal outlier status (three standard deviations from the mean). To explore the resulting findings from dual statistical perspectives, we used both the Pearson correlation coefficient and its Bayesian equivalent. The Bayesian analysis was particularly important for evaluating associations between fMRI signals and EEG HEP signals as it facilitated the testing of evidence for the null hypothesis (negative log(BF_10_) values, Supplementary Methods 5; Table S2). It was performed in JASP version 0.14.1 (https://jasp-stats.org/).

## RESULTS

Of the 48 female study participants with analyzable EEG data, 24 were individuals with GAD and 24 were matched HCs based on self-reported age and measured BMI (Table 1; Figure S1).

**Table 1.** Summary of demographic and clinical measures for general anxiety disorder (GAD) and healthy comparison (HC) participants.

| Variable | GAD<br>(n=24 females) | HC<br>(n=24 females) | Statistic |  |
| --- | --- | --- | --- | --- |
|  | M [SD] |  | P-value | Log(BF<sub>10</sub>) |
| Age (years) | 26.5 [6.9] | 24.3 [5.6] | 0.41 | -1.09 |
| BMI (kg/m<sup>2</sup>) | 25.9 [4.9] | 24.1 [3.4] | 0.27 | -0.74 |
| GAD-7 | 13.8 [3.2] | 1.0 [1.5] | <0.001 | 7.71 |
| OASIS | 10.8 [2.1] | 1.4 [1.7] | <0.001 | 6.97 |
| PHQ-9 | 10.8 [5.2] | 1.0 [1.1] | <0.001 | 7.92 |
| ASI-Physical | 6.3 [5.0] | 1.6 [1.8] | <0.001 | 2.47 |

Notes. M: mean; SD: standard deviation; Log(BF<sub>10</sub>): log scale of Bayes factor BF10; BMI: Body Mass Index; kg: kilograms; m: meters; GAD-7: 7-item Generalized Anxiety Disorder scale; OASIS: Overall Anxiety Severity and Impairment Scale; PHQ-9, 9-item Patient Health; PHQ-9: 9-item Patient Health Questionnaire; ASI-Physical: Physical Concerns subscale of the Anxiety Severity and Impairment Scale

### HEP changes between saline and isoproterenol 0.5 μg

#### Peak period

We observed a significant group effect over midline right frontocentral and parietal electrodes when comparing the change in HEP amplitude between the saline and the 0.5 μg infusions during the peak period. This effect was observed within a 44 millisecond (ms) window from 228 to 272 ms after the cardiac R-wave event (Monte-Carlo P=0.009, Cohen’s *d*=0.18; Figure 1A, left panel; Table S3), and included the following electrodes: F4, C4, P4, T8, P8, FC2, CP2, FC6, CP6, and TP10. Additionally, there was an opposing effect across groups on the changes in HEP amplitude between conditions. Specifically, the GAD group had positive HEP amplitude changes from the saline to the 0.5 μg infusions whereas HCs had negative HEP amplitude changes (Figure 1A, right panels). Within-group analysis indicated that the positive change in HEP amplitude between the saline and the 0.5 μg infusions in the GAD group was significant from 252 to 276 ms after the cardiac R-wave event (Monte-Carlo P=0.02, Cohen’s d=0.54), whereas the corresponding HEP amplitude change in the HC group was negative and did not reach the significance threshold (Monte-Carlo P=0.30, Cohen’s d=0.24).

**Figure 1.**
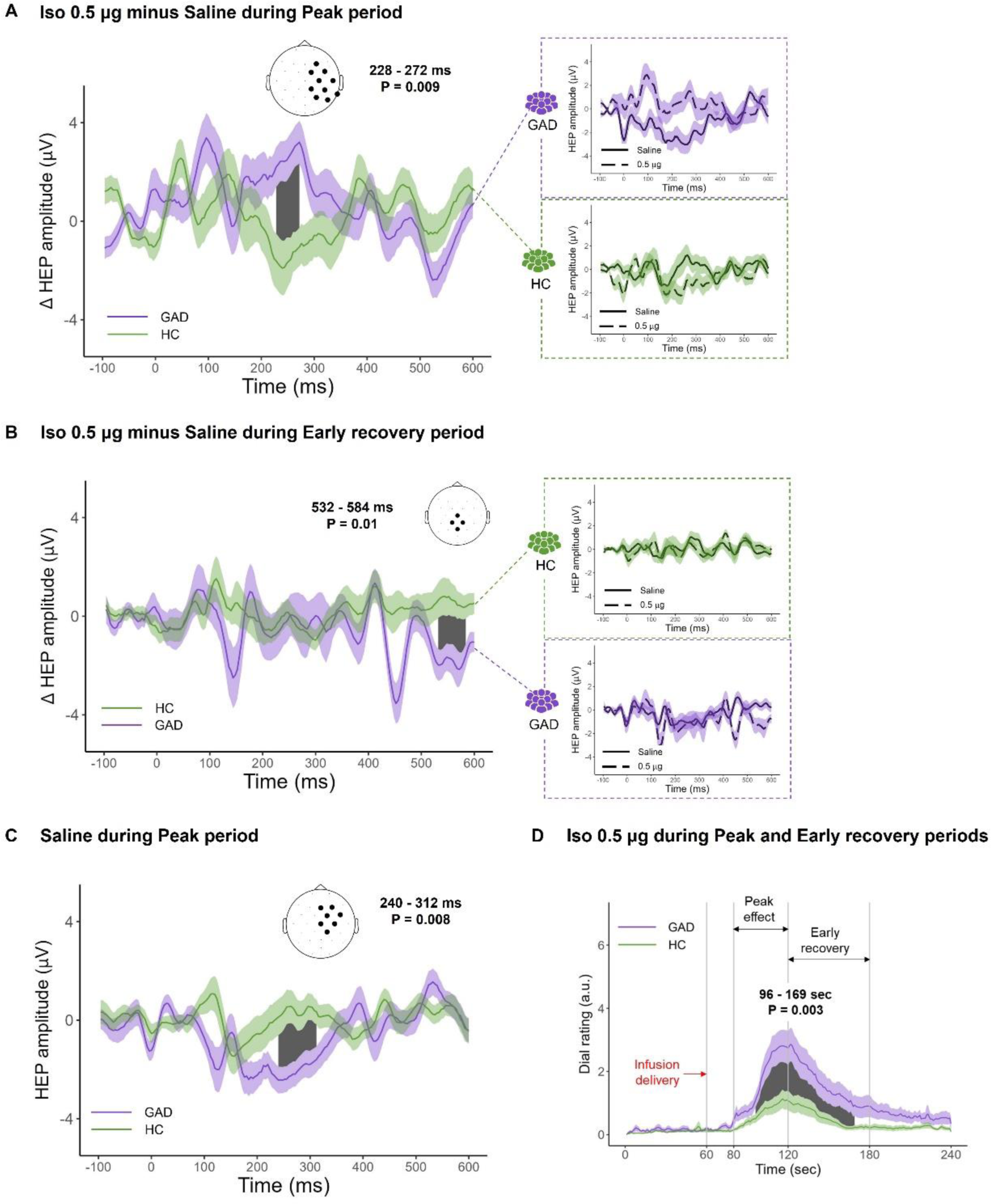
Isoproterenol-induced changes in HEP amplitude and cardiorespiratory sensation. **(A, left)** Greater isoproterenol-induced changes in HEP amplitude for GAD versus HC in right frontocentral and parietal electrodes during the peak period. A significant group effect was found when analyzing the change in HEP amplitude between the peak period of the 0.5 μg and saline infusions from 228 to 272 ms. Black dots on the scalp map indicate the EEG electrodes contributing to the significant cluster time window (indicated by the grey shaded area) whose limits are indicated in milliseconds (ms) from the cardiac R-wave event (occurring at 0 ms on the x axis). Grand-averaged HEP waveforms were computed from the electrodes identified in the significant cluster. **(A, right)** Opposing shifts in HEP amplitude between the saline and the 0.5 μg infusions for the GAD and HC groups. The GAD group showed more positive HEP amplitudes during the 0.5 μg infusions whereas the HC group showed more negative HEP amplitudes. *Note: the grand-averaged HEP waveforms in (A, right) were computed across the electrodes identified in the significant cluster reported in (A, left).* **(B, left)** A significant group effect (GAD vs HC) was found when analyzing the change in HEP amplitude between the saline and the 0.5 μg infusions during the early recovery period, from 532 to 584 ms after the cardiac R-wave event. **(B, right)** Shifts in HEP amplitude between the saline and the 0.5 μg infusions for the GAD and HC groups separately. **(C)** During the peak period of saline infusion, the GAD group showed significantly greater HEP amplitude than the HC group in right frontocentral electrodes from 240 to 312 ms after the cardiac R-wave event. **(D)** Greater isoproterenol-induced cardiorespiratory sensation in GAD versus HC during the 0.5 μg infusion. Waveforms represent the average of cardiorespiratory sensation intensity that was continuously rated by participants using a dial ranging from 0 (none or normal) to 10 (most ever). The grey shaded area highlights the window where a significant difference was observed between GAD and HC. GAD – generalized anxiety disorder, HC – healthy comparison, Iso – isoproterenol, μg – micrograms, μV - microvolts, ms – milliseconds, sec - seconds, a.u. – arbitrary unit. Error bars indicate 95% confidence intervals.

#### Early recovery period

We found significant group differences in HEP amplitude changes between the saline and the 0.5 μg infusions during the early recovery period. The GAD group had significantly larger changes in HEP amplitude compared with HCs at midline frontocentral, central, and centroparietal electrodes. The group effect was observed within a 52 ms window from 532 to 584 ms after the cardiac R-wave event (Monte-Carlo P=0.01, Cohen’s d=0.44; Figure 1B; Table S3), and included the Cz, Pz, CP1, and CP2 electrodes.

### HEP changes between saline and isoproterenol 2 μg

No group differences were found for changes in HEP amplitude between the saline and the 2.0 μg infusions during the peak or the early recovery periods (Table 2; Figure S7).

**Table 2.** Statistics from cluster based permutation analysis informing on how HEP amplitudes differed between the GAD and HC groups (n=24 each), based on comparisons made within and between conditions.

|  | *Periods* | *Peak* |  | *Early recovery* |  |
| --- | --- | --- | --- | --- | --- |
|  |  | *P* | *d* | *P* | *d* |
| *Within condition* |  |  |  |  |  |
| **Saline** |  | 0.008 | 0.46 | 0.007 | 0.28 |
| **Iso 0.5 µg** |  | 0.12 | 0.47 | 0.17 | 0.84 |
| **Iso 2 µg** |  | 0.18 | 0.18 | 0.26 | 0.29 |
| *Changes between conditions* |  |  |  |  |  |
| **Saline vs. Iso 0.5 µg** |  | 0.009 | 0.18 | 0.01 | 0.44 |
| **Saline vs. Iso 2 µg** |  | 0.29 | 0.37 | *nc* | *nc* |
| **Iso 0.5 µg vs. Iso 2 µg** |  | 0.25 | 0.10 | 0.11 | 0.30 |

Notes. GAD: generalized anxiety disorder; HC: healthy comparison; HEP: heartbeat evoked potential; P: Monte-Carlo p-value; d: Cohen's d for effect size; Iso: isoproterenol; µg: micrograms; nc: no cluster identified at the first step of cluster permutation analysis, so no evaluation of statistical significance under permutation distribution

### HEP during saline condition

#### Peak period

The GAD group had significantly larger HEP amplitudes (in absolute value) than HCs over right frontocentral electrodes during the peak period of saline infusion. The effect was observed within a 72 ms window from 240 to 312ms after the cardiac R-wave event (Monte-Carlo P=0.008, Cohen’s *d*=0.46; Figure 1C; Table S3), and including the Fz, F4, Cz, C4, FC2, FC6, and CP2 electrodes.

#### Early recovery period

We observed a similar group effect during the early recovery period of saline infusion. The GAD group showed significantly larger HEP amplitudes (in absolute value) than HCs at right frontocentral and parietal electrodes (including the F4, C4, P4, Cz, Fz, FC2, CP2, and FC6 electrodes), within a 72 ms window from 240 to 312 ms after the cardiac R-wave event (Monte-Carlo P=0.007, Cohen’s d=0.28).

### HEP during Isoproterenol 0.5 μg and 2 μg conditions

No significant group differences were found during the peak and early recovery periods of the 0.5 μg and 2.0 μg doses of isoproterenol.

Table 2 summaries the data on how HEP amplitudes differed between the GAD and HC groups, based on various comparisons made both within and between conditions.

### Interoceptive awareness

The GAD group reported a significantly greater intensity of cardiorespiratory sensations than HCs during the 0.5 μg dose of isoproterenol from 36 to 109 seconds after the onset of infusion delivery (Monte-Carlo P=0.003, Cohen’s d=0.78; Figure 1D), corresponding to an epoch spanning the peak and early recovery periods. However, there were no significant group differences in heart rate across any dose (Figure S8), and no differences in cardiorespiratory sensation ratings during the saline and the 2.0 μg infusions (Figure S9).

### Multilevel Correlation Analyses

No correlations across participants emerged between average HEP amplitude (either for saline alone and the change between saline and 0.5 infusion conditions) and: (1) average self-reported cardiorespiratory intensity (Figure S10, upper panel); (2) self-reported anxiety (Figure S10, lower panels); (3) heart rate (Figure S11); or (4) fMRI signal in regions of interest (insula, vmPFC, dACC, amygdala, or somatosensory cortex) in the subset of 21 GAD and 21 HC participants with usable HEP-fMRI data (Table 3). However, the average HEP amplitude calculated from the peak period HEP cluster of the saline infusion was negatively correlated with PHQ-9 depression (*r*=-0.37, *p*=0.01 uncorrected, log(BF_10_)=1.49) and ASI physical concerns (*r*=-0.35, *p*=0.02 uncorrected, log(BF_10_)=1.17) (Figure 2).

**Figure 2.**
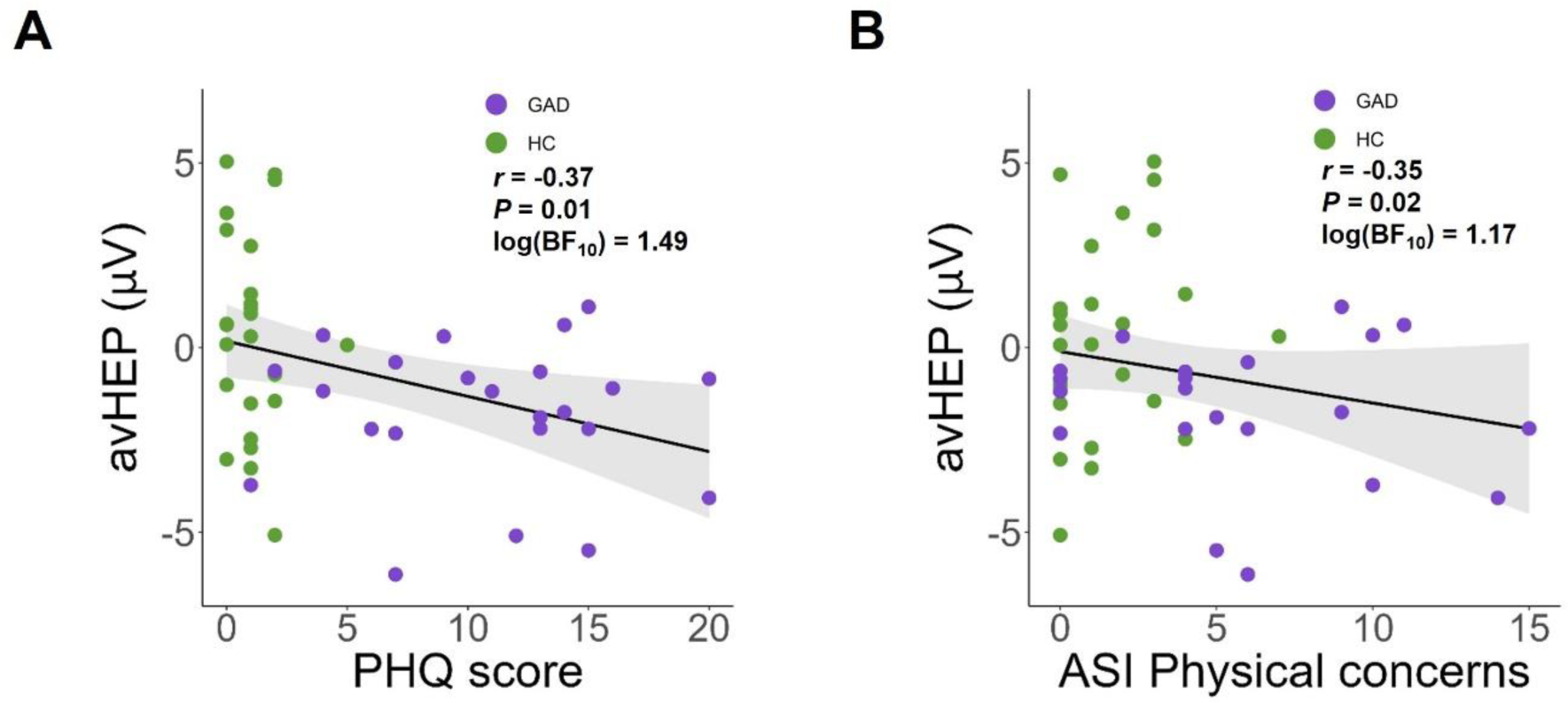
Association between two self-reported personality traits and the average HEP amplitude (avHEP) during the peak period of the saline infusion: **(A)** the 9-item Patient Health Questionnaire (PHQ) for depression, and **(B)** the Physical Concerns subscale of the Anxiety Severity and Impairment (ASI) Scale. These associations show that the higher the depression score for the PHQ-9 and the ASI physical concerns, the more negative the avHEP value. Note that the correlations between the avHEP and the PHQ-9 and ASI physical concerns scores are negative because of the negative polarity of HEP signal during saline infusion (see Figure 1C). As these correlations are exploratory and primarily intended to be descriptive, they are not corrected for multiple comparisons; Bayes factors are also included. μV – microvolts

**Table 3.** Correlations (2-tailed) between recorded HEP signals and BOLD fMRI signals in a subsample of 21 GAD and 21 HCs with both EEG and fMRI signals available.

| HEP signal | fMRI signal | Statistic |  |  |
| --- | --- | --- | --- | --- |
|  |  | r | p | Log(BF<sub>10</sub>) |
| avHEP<sup>#</sup><br>during Peak period<br>of the saline infusion | Insula | -0.08 | 0.62 | -1.53 |
|  | vmPFC | 0.02 | 0.88 | -1.64 |
|  | dACC | -0.05 | 0.73 | -1.59 |
|  | Somatosensory cortex | 0.08 | 0.62 | -1.53 |
|  | Amygdala | 0.04 | 0.79 | -1.61 |
| avHEP<sup>#</sup><br>Iso 0.5 µg minus Saline<br>during Peak period | Insula | -0.03 | 0.84 | -1.64 |
|  | vmPFC | -0.26 | 0.09 | -0.29 |
|  | dACC | -0.12 | 0.42 | -1.40 |
|  | Somatosensory cortex | 0.23 | 0.14 | -0.57 |
|  | Amygdala | -0.09 | 0.56 | -1.50 |

Notes. # - average HEP signal across all electrodes and time points that were included in the corresponding HEP cluster showing significant group differences (HC vs. GAD), vmPFC - ventromedial prefrontal cortex, dACC - dorsal anterior cingulate cortex, µg – micrograms, r – Pearson correlation coefficient, p - p-value, Log(BF<sub>10</sub>) - log scale of Bayes factor BF<sub>10</sub>. Correlations are intended to be descriptive and are not corrected for multiple comparisons.

## DISCUSSION

In this study investigating heartbeat evoked potentials (HEP) in females with generalized anxiety disorder and age-, sex-, and BMI-matched healthy comparisons (HC), we observed significant group differences in HEP amplitude changes between isoproterenol-induced peripheral adrenergic stimulation and saline infusion. The GAD group showed more positive HEP amplitude changes between the saline and 0.5 μg dose of isoproterenol during the peak period of infusions and more negative amplitude changes during the early recovery period, compared to HCs. Additionally, the GAD group exhibited larger HEP amplitudes in absolute value during saline infusions, when cardiovascular tone did not increase. HEP amplitudes during saline infusions were also negatively associated with depression scores on the PHQ-9 and the Physical Concerns subscale of the ASI across both groups. However, no significant association was found between HEP and BOLD fMRI signals in the insula, vmPFC, dACC, amygdala, or somatosensory cortex.

Our HEP findings indicate that some fundamental electrophysiological differences exist between individuals with GAD and HCs during low levels of peripheral adrenergic stimulation. During the peak period, participants with GAD demonstrated larger HEP amplitude changes compared to HCs, with a latency of this effect that was similar to the meta-analytic effects of arousal reported previously (i.e., between 200 to 300 ms after the R-wave [17]). The right frontocentral and parietal electrode location of this effect was similar to electrode regions engaged during high arousal conditions in previous HEP studies, such as the larger HEP amplitudes induced during affective auditory or visual cue presentation [21], or by the intravenous infusion of 4milligrams of the adrenal hormone cortisol [22] in healthy individuals. The HEP differences at the 0.5μg dose of isoproterenol were also accompanied by heightened interoceptive awareness in the GAD group, yet there was no substantial group difference in heart rate. Taken together, these empirical findings provide evidence in favor of a central autonomic hypersensitivity in GAD. While we recognize the complexities in interpreting variations in HEP magnitude and directional shifts, it is important to clarify that these variations do not directly inform whether the observed neural activity is excitatory or inhibitory. Given the prior literature suggesting that the HEP is a marker for cortical processing of heartbeats [17], one might expect that individuals with GAD would show increased HEP magnitudes during the infusion of 0.5μg isoproterenol as compared to saline, as a sign of autonomic hyperarousal. Contrary to this expectation, however, our findings revealed that the HEP amplitude was actually smaller in the GAD group during the 0.5 μg isoproterenol infusion compared to the saline condition. While it could be tempting to interpret this finding as indicative of decreased cortical neural activity in individuals with GAD, such an interpretation would be speculative. If the HEP amplitude was to reflect the amount of cortical resources allocated to processing a condition, the smaller HEP during 0.5μg isoproterenol versus saline within the GAD group could be interpreted to reflect less effortful processing, given that they likely expected greater isoproterenol-induced cardiac change. Thus, our study does not offer a straightforward explanation for how variations in HEP magnitude and directionality relate to individual differences in cardiac interoception in GAD.

Moreover, it’s worth noting that differences in HEP amplitude may not necessarily be tied to interoceptive processes. Some research suggests that HEP amplitudes could reflect variations in cardiac dynamics such as stroke volume or arousal [21, 44], rather than differences in the brain’s processing of bodily sensations. The influence of cardiac dynamics on HEP amplitude was one of the factors of interest in the present work. Indeed, previous HEP studies focusing on clinical groups, including individuals with GAD, have only investigated the HEP measure under conditions of physiological rest or during sleep [17]. The goal of this previous work was to characterize differences in HEP amplitude between clinical and healthy samples, while assuming a lack of differences in their peripheral physiology. In this study, we directly assessed the impact of acute changes in cardiovascular state on HEP amplitude in order to elucidate the abnormal physiological arousal implicated in GAD pathophysiology [4, 5], using experimentally-induced adrenergic stimulation. Our study offers nuanced insights into the electrophysiological differences between individuals with GAD and healthy individuals, particularly in response to low-level peripheral adrenergic stimulation. While our findings suggest central autonomic hypersensitivity in GAD, they underscore the complexity of interpreting HEP measures, which may be influenced by both neural and cardiac dynamics. This work advances our understanding of the role of abnormal physiological arousal in GAD pathophysiology, emphasizing the need for further research to disentangle the neural and cardiac contributions to these observed differences.

Regarding the lack of group differences in HEP amplitude during the 2μg isoproterenol condition, we believe that it could reflect a ceiling effect of the isoproterenol-mediated approach. We observed a similar lack of group effect for other measures of cardiac interoception, such as the fMRI signals in the insular cortex and vmPFC, as previously reported [15], suggesting that the 2μg dose is sufficient to exert a global increase in the processing of cardiovascular signals irrespective of clinical condition. We do not believe interoceptive sensitivity in GAD would manifest only at lower levels of adrenergic stimulation, and consistent with this perspective, the GAD patients reported greater anxiety at both the 0.5 and 2μg doses in our previous study, with no differences in response to saline [15]. However, it has been suggested that dysfunctional interoception in GAD affects perceptual discriminative mechanisms which underly one’s ability to discriminate between two interoceptive signals characterized with different signal-to-noise ratios [9, 27]. In our study, the group differences at low level (0.5μg) but not at high level (2μg) of adrenergic stimulation could be interpreted as evidence in favor of this hypothesis, because the signal-to-noise ratio of cardiac signals can be described as weak during low-level adrenergic stimulation (corroborated by a relatively limited increase in heart rate), contrary to high-level adrenergic stimulation that increases the signal-to-noise ratio of cardiac signals through substantial increases in heart rate and contractility. Determining whether increased interoceptive sensitivity in GAD manifests only at lower levels of peripheral adrenergic stimulation would require studies that manipulated additional levels of low and high arousal in GAD (e.g., 0.75μg and 1.0μg doses, as we have used previously [12, 14]). One could adopt a similar approach to evaluating respiratory interoception using different inspiratory breathing load perturbations (e.g. ranging from 5 to 40 cmH_2_O/L/sec inspiratory resistance loads (as in [45]), or higher).

The mechanistic basis for central autonomic hypersensitivity in GAD could involve both bottom-up and top-down processes associated with abnormal interoceptive processing. Double-blinded administrations of isoproterenol and saline allowed us to experimentally manipulate participant’s expectancies about infusion-elicited bodily changes, and the larger HEP amplitudes in absolute value observed during saline infusions in participants with GAD might be due to abnormalities in top-down interoceptive processing. Since infusion-elicited changes in cardiac sensations (including increased heart rate and heart palpitations) were most likely to occur during the initial minute following the delivery an infusion, the lack of arrival of such changes during saline infusion could be presumed to result in a mismatch between interoceptive predictions and actual interoceptive sensory inputs. The HEP amplitude has been proposed to reflect this interoceptive prediction error, based on the fact that top-down attention towards the heart, which increases prediction error by improving precision of cardiac input, modulates HEP amplitude [24], and the fact that prediction errors are thought to be encoded by superficial pyramidal cells that are the major contributor to empirically observed neurophysiological responses [23, 46]. If the females with GAD had stronger expectations about situations eliciting bodily changes leading to a larger mismatch between expectations and actual cardiovascular states during isoproterenol stimulation, this could result in an increased HEP amplitude, reflecting a heightened interoceptive prediction error (see Supplementary Discussion). This interpretation does not require that interoceptive prediction errors to be conscious phenomena, as they can occur even in the absence of group differences in conscious sensation [47]. It should be noted that a complete separation of the bottom-up and top-down components of cardiovascular signal processing remains challenging from an experimental standpoint. In our protocol, participants were informed (i.e., their expectations were cued) at the onset of each infusion, meaning that top-down processing was engaged across all conditions even though they were blinded regarding the compound (isoproterenol vs. saline) being administered. However, we did not include other conditions in which top-down processing was not engaged (e.g., no cue to attend to cardiorespiratory sensations during covert isoproterenol administration or during no infusion at all). Incorporating these additional comparisons in future studies would be helpful to fully disentangle the influence of the top-down from bottom-up components of interoception.

HEP amplitudes did not correlate with any acute physiological or subjective responses, including HR, anxiety, and self-reported intensity ratings of cardiorespiratory sensation, but they did correlate with some trait measures of symptomatology (i.e., depression and anxiety sensitivity physical concerns). The involvement of physical and mood-related concerns adds an affective dimension to the interpretation of the HEP, emphasizing that both sensory and symptom characteristics should be considered when evaluating factors that modulate HEP responses. The lack of correlation between HEP and fMRI signals in the insula, vmPFC, dACC, amygdala, and somatosensory cortex also suggests the need for further investigation into the neural sources responsible for the HEP signal in GAD. It should be noted that intrinsic differences between HEP and fMRI signals in terms of time resolution (millisecond for HEP versus second for fMRI) may only partly explain the statistical independence that we observed between fluctuations of HEP amplitude and fMRI signals. Other methodologic limitations could also have contributed. For example, from an event-related standpoint, the low number of isoproterenol infusion trials may have reduced the sensitivity of the fMRI measures to measure BOLD responses from certain regions. Such an analysis would support a whole-brain analysis of covariation between the BOLD and EEG signals, but it would also require a much larger number of infusions, which could impose a challenge to the feasibility this approach within the fMRI environment. Therefore, the present lack of correlation between the HEP response differences and fMRI differences is not definitive proof of a lack of involvement of the brain regions that we have examined (the insula, vmPFC, dACC, amygdala, and somatosensory cortex). Overall, these null findings suggest that fully elucidating the specific neural mechanisms and pathways underlying cardiac interoceptive dysfunction in GAD will require additional approaches that account for the dual influences of bottom-up and top-down mechanisms on the neural processing of cardiac signals.

### Limitations

This study focused exclusively on females with GAD due to the fact that females outnumber males on a 2:1 basis [48]. Although previous HEP studies do not suggest a gender effect on HEP amplitude [17], future research will need to establish whether the current findings may extend to males with GAD. We computed the HEP measure from EEG data that was simultaneously recorded during fMRI signal acquisition, necessitating the careful removal of complex artifacts such as the BCG and CFA artifacts [49-52]. This represented a methodological challenge for HEP computation because such signals are intrinsically amplified when synchronizing and averaging EEG signal on cardiac R-wave event [53-56]. In the present work, by combining the OBS and ICA approaches we were able to successfully remove these complex artifacts from the EEG signal. However, it is possible that our processing approach obscured some EEG signals relevant to the HEP (e.g., either because of overly aggressive data cleaning to remove fMRI- or cardiac-related artifacts, due to an obscuring of the signal by artifact residuals, or due to persistent influences of isoproterenol-induced cardiac output changes on heartbeat evoked responses [44]). We can argue against this possibility by virtue of the fact that we observed relevant heartbeat evoked responses in the EEG signal, in both systole- and diastole-related components and in electrode locations that are consistent with those identified in prior HEP studies [17].

## CONCLUSION

This study provides evidence for distinct electrophysiological responses to heartbeats in females with GAD compared to age-, sex-, and BMI-matched HCs during peripheral adrenergic stimulation. The findings support a central autonomic hypersensitivity in GAD, characterized by HEP profiles that are intricately tied to both bottom-up and top-down interoceptive processes, as well as to trait measures of symptomatology. Our analysis does not establish a direct correlation between HEP and fMRI signals in brain regions known as key interoceptive hubs. Beyond the intrinsic differences between the two signals, the statistical independence of electrophysiological signals from blood oxygen level-dependent neural responses suggests that there might be multiple mechanisms of cardiac interoceptive dysfunction in GAD, warranting further investigations focused on evaluating the complex interplay of neural and cardiac dynamics in this disorder.

## Data Availability

The data and materials are currently private for peer review.

## Acknowledgments.

Megan Sinik, B.S., Dhvanit Raval, B.S., and Chloe Sigman, B.S., provided physiological data processing assistance. Rachel Lapidus Ph.D., Valerie Upshaw, M.S.N., C.N.A.P., and Maria Puhl, Ph.D., provided data collection assistance. AMegan Cole, R.N., Jeanne Echols, R.N., Lisa Augustine, R.N., Susan Maxey, R.N., and Lindsey Bailey, N.T., provided isoproterenol preparation assistance. We also acknowledge the key contributions of the late Jerzy Bodurka to the acquisition of the functional neuroimaging data. The results included in the present paper were presented at the American College of Neuropsychopharmacology meeting, in December 2022.

## Author Contributions

Concept and design: Khalsa. Acquisition, analysis, or interpretation of data: Verdonk, Teed, White, Ren, Stewart, Paulus, Khalsa. Drafting of the manuscript: Verdonk, Khalsa. Critical revision of the manuscript for important intellectual content: Verdonk, Teed, White, Ren, Stewart, Paulus, Khalsa. Statistical analysis: Verdonk, Khalsa. Obtained funding: Paulus, Khalsa. Administrative, technical, or material support: Khalsa. Supervision: Khalsa.

## SUPPLEMENTARY CONTENT

**Supplementary Methods 1.** Participants

One hundred thirty-six participants were assessed for eligibility. 11 candidates declined to participate and the remaining 125 provided informed written consent. 17 participants withdrew prior to the intervention for various reasons (scheduling difficulty, incomplete baseline assessment, or study refusal). Thirty-five additional participants were excluded due to contraindicated medical conditions, including current or prior cardiac disease. Of the 73 participants allocated to the intervention, 72 completed it, including 36 individuals with generalized anxiety disorder (GAD). One GAD participant withdrew during the infusions due to panic anxiety. After completion of the intervention, data related to 11 GAD participants were discarded: four due to equipment failures; three due to issues concerning the identification of experimental condition in data files; and four due to excessive head motion resulting in low EEG data quality (Figure S1). The data were collected from January 1, 2017, to November 31, 2019, at the Laureate Institute for Brain research (Tulsa, Oklahoma).

**Supplementary Methods 2.** Experimental protocol

Isoproterenol was obtained from Valeant Pharmaceuticals (Laval, QC, Canada). Infusions were prepared into 3 mL boluses by a pharmacist who was unblinded. The isoproterenol infusion administration procedure mirrored our previous fMRI cross-over protocol in healthy volunteers [1], with infusions administered by a nurse seated beside the scanning bed inside of the MRI room who was blinded to infusion condition. For safety monitoring purposes, cardiac rhythm was also recorded continuously using two MRI-compatible ECG leads (lead I and II configuration) (GE Healthcare, Waukesha, WI, USA). These rhythms and vital signs were monitored continuously by the nurse delivering the infusions. As an additional safety precaution, condition order was available to the research assistant sitting in the control room, who was blinded to the study hypotheses.

The randomized sequences for dose order were individually predetermined via a random number generator prior to beginning study sample recruitment. Upon recruitment of each participant, a 3rd party registered nurse (i.e., uninvolved in infusion administration or data collection) selected a randomization code from a set of randomizations generated by the senior study author at the beginning of the study. The 3rd party nurse arranged the bolus doses in the predetermined order, and after obtaining verification of this order by a second 3rd party nurse (or nursing assistant), covered the labels. Selection of a randomization order for a given participant was not determined by group membership and occurred in sequential order from the randomization list.

On arrival participants ate a 300 Calorie snack. Each participant was led through a training session approximately one hour before fMRI scanning. During this session, the participant was instructed that they would receive both isoproterenol and saline infusions at different points during the scan. Participants were informed that “you may notice an increase in your heartbeat sensations, and/or may notice increase in your breathing sensations” during the isoproterenol infusions. To familiarize participants with the experience of the infusion setup and isoproterenol-induced sensations, prior to the scan, participants received two practice bolus infusions (saline and 1 microgram, *μ*g) delivered by the nurse in a separate room near the MRI scanning suite. These infusions followed the same time course as those administered during the scanning session and required participants to provide sensation ratings using a dial. We administered a 1 *μ*g dose during the practice infusion, to avoid a familiarity effect during the subsequent infusion scans and to ensure that participants received a large enough dose that they were likely to perceive based on our prior studies [1-3]. Following the scan session participants ate a 1000 Calorie meal.

**Supplementary Methods 3.** Computation of the heartbeat evoked potential The recorded raw EEG-ECG data were imported to EEGLAB, then we processed them using custom scripts in Matlab 2020b (Mathworks^®^) and the EEGLAB toolbox (version 19.0; [5]) for artifact correction, downsampling, bandpass filtering, and re-referencing. The fMRI-related gradient artifact was removed from the EEG and ECG signals using the fMRIb plug-in (version 2.00) for EEGLAB, specifically the command *pop_fmrib_fastr* that implements the Optimal Basis Set (OBS) method (Figure S3).

The cardiac-related ballistocardiogram (BCG) artifact was removed using the OBS method [4], specifically the command *pop_fmrib_pas* of the fMRIb plug-in for EEGLAB (Figure S4).

Independent Component Analysis (ICA) was used to correct for eye-related artifacts, including eyeblinks and saccadic eye movements. Independent components (ICs) were identified as related to ocular artifacts based on the time course of the signal, topographic map, power spectrum density, and energy. We used decision criteria of two validated approaches for artifact removal from EEG data acquired simultaneously with fMRI. First, we used the automatic EEG-assisted retrospective motion correction for fMRI (aE-REMCOR) approach [5], which was developed specifically to remove eyeblink and other EEG artifacts in the fMRI environment. This involves the application of decision criteria that are based on the spectrum features and the topographic maps of ICs (e.g., see Tables S3 and S5 in Supplement of [5]). The second method, the rtICA approach [6] involves decision criteria that are based on the spectral characteristics (which differ from the aE-REMCOR approach), the energy, and kurtosis features (see Table 2 of [6]).

Data were epoched with the cardiac R-wave event as the temporal reference, from -100 ms to 650 ms after R-wave. To facilitate this, we used additional information from the PPG waveform to improve cardiac R-wave event detection as ECG signal quality can be deteriorated by gradient artifact correction in some individuals. Based on previous findings showing a delay of approximately 250 ms between the ECG R-peak and the peak of the finger PPG waveform [7, 8], a custom Matlab script was applied to detect R-peaks on the ECG signal within a time window from -350 to -150 milliseconds (ms) from the peak of the finger PPG waveform (Figure S5). Afterwards, the ECG data for each participant were visually inspected to assure that the cardiac R wave events were correctly identified.

We implemented on epoched data an additional step for correcting potential BCG residuals using ICA. Based on the decision criteria proposed by Debener et al (2008), BCG residual-related ICs were identified by the amount of variance they contributed to the evoked pulse artifact, using the EEGLAB function *eeg_pvaf()* [9, 10]. Specifically, ICs accounting for higher than 5% of the variance of the evoked signal (including EEG and ECG signals) in the time range [0-650 ms] after the cardiac R-wave event were removed (Figure S6A). The ICA method was also used to correct the cardiac field artifact (CFA) by removing ICs whose amplitude was higher than 5 μV within the time window from -100 to 100 ms from the cardiac R-wave event, and whose timing over heart cycles resembled the characteristics of the CFA (Figure S6B) [11]. On average, there were 7 ICs that were removed per participant per condition (Supplementary Results 2).

**Supplementary Methods 4.** MRI data acquisition, preprocessing, and analysis

MRI scans were acquired in a GE MR750 3T scanner with an 8-channel head coil allowing for simultaneous EEG recordings. Anatomical images were acquired via a T1-weighted magnetization-prepared rapid gradient echo (MPRAGE) sequence with sensitivity encoding (SENSE11) that lasted 5 min and 40 s. MPRAGE parameters were: 190 axial slices, slice thickness = 0.9 mm, TR/TE = 5/2.012 ms, FOV = 240 x 192 mm2, matrix size 256 x 256, flip angle = 80, inversion time = 725 ms, SENSE acceleration R = 2, with a sampling bandwidth of 31.2. A 240 s, single-shot gradient echo planar imaging (EPI) sequence covering the whole-brain was used for each fMRI scan. In this EPI sequence we obtained axial 39 slices, 2.9 mm thick, with no gap and a voxel size of 1.875 x 1.875 x 2.9 mm3. Additional parameters were TR/TE = 2000/27 ms, FOV = 240 x 240 mm2, flip angle = 780, SENSE acceleration R = 2 with a 96 x 96 matrix.

The first 4 EPI volumes were dropped to allow field stabilization. BOLD signal was scaled to percent change from the time-course mean for each voxel. With respect to motion artifacts, poor quality volumes were censored using interpolation if they exceeded 0.3 mm of mean motion or if >20% of voxels were found to be outliers using AFNI’s *3dtoutcount*. Runs were discarded if >20% of volumes were censored.

A whole-brain analysis was conducted using AFNI’s *3dttest*++ program and applied the ETAC (Equitable Thresholding and Clustering) option to estimate significant cluster sizes corresponding to a 5% false positive rate at a *p* < 0.001 voxel-wise threshold [12]. We selected this method to reduce arbitrary judgment in selection of the uncorrected p-value threshold [13].

**Supplementary Methods 5.** Statistical analysis

The main idea behind the cluster permutation approach is that, if an effect is robust and biologically relevant, it will be present in neighbouring electrodes and time points. In addition, the method tests the significance of the effect by comparing the actual statistical result against a distribution of the statistical values obtained by comparing the same data points after shuffling [14, 15]. We considered this exploratory statistical approach most suitable for the present study due to the considerable variability of HEP features that have been reported [16] and due to our lack of prior assumptions about the topography and the latency of a potential effect of diagnostic group on HEP amplitude.

The following steps were taken to identify clusters: (a) t-statistics were computed between diagnostic groups (GAD vs HC) for each samples in the spatio-temporal HEP data; (b) these sample-specific statistics were thresholded by p-value (p < 0.05); (c) neighbouring data points were identified that exceeded the threshold and had the same sign; (d) cluster-level statistics were calculated by summing the t-statistics; and (f) this maximum was evaluated under its permutation distribution. The permutation distribution was derived from the statistical values of independent t-tests based on 10,000 random permutations of HEP data with respect to diagnostic group, and the p-value threshold for the inclusion in the cluster was set at 0.05 [17]. The cluster permutation was run considering the spatial (electrodes) and temporal dimensions (time-points) of the dataset. Electrodes less than 2.5 cm apart were considered neighbours, resulting in an average of 5.5 neighbours per electrode. The minimum number of neighborhood electrodes that was required for a selected sample to be included in the clustering algorithm was three electrodes. We considered a significant cluster to be biologically relevant if it was observed for a period of at least 30 ms, i.e., capable of supporting a train of neocortical action potentials whose duration is typically 2 ms [18], and consistent with minimum durations reported in the meta-analytic HEP literature [16]. For the purpose of statistical analysis, we performed a final correction on the grand-averaged HEP waveform (i.e. at the group level) by subtracting the mean of the 100 ms preceding the time window of interest (i.e., from 0 to 100 ms after the R-wave) from the entire signal, in order to prevent any residual influence of the CFA on HEP signal.

We also used the non-parametric cluster permutation approach to evaluate isoproterenol’s effects the time course of heart rate (HR), self-reported cardiorespiratory intensity, and the ECG waveform (the latter to test for direct effects on the cardiac waveform). This analysis was performed between diagnostic groups (i.e., GAD vs HC) at each time point of the 240 seconds period of infusion, for the HR and the self-reported cardiorespiratory intensity. For the ECG waveform, the cluster permutation analysis was performed in the same time range as for the HEP waveform, namely from 100 to 600 ms after the cardiac R-wave event.

The relationship between neural, cardiac, and subjective outcomes was examined through correlation analysis. To gain a better understanding of the results, we used both the Pearson correlation coefficient and its Bayesian equivalent. We calculated the log scale of the Bayes factor (*log(BF_10_)*) to determine the likelihood of the alternative hypothesis compared with the null hypothesis. The *log(BF_10_)* value can be easily interpreted such that a negative value indicates support for the null hypothesis, whereas a positive value indicates evidence in favor of the alternative hypothesis (see Table S2 for an interpretation scale of *log(BF_10_)*).

**Supplementary Results 1.** ERP analysis

The ERP analysis included 435 epochs (Standard Deviation, SD: 116) per participant during the saline condition, 467 epochs (SD: 116) during the 0.5-μg dose, and 515 epochs (SD: 107) during the 2-μg dose. The number of included epochs did not differ significantly between the groups during the 0.5-μg (W=284, P=0.94, log(BF_10_)=-1.18; GAD: mean: 470, SD: 101; HC: mean: 463, SD: 132) or 2-μg doses of isoproterenol (W=265, P=0.64, log(BF_10_)=-1.17; GAD: mean: 526, SD: 91; HC: mean: 506, SD: 123). During the saline condition, the number of included epochs was marginally lower in the GAD group compared to HC (W=193, P=0.05, log(BF_10_)=0.89; GAD: mean: 396, SD: 120; HC: mean: 475, SD: 100).

**Supplementary Results 2.** Independent components (ICs) identified as related to cardiac-specific artifacts using Independent Component Analysis (ICA)

In total and on average, the ICA approach identified 6 ICs (SD: 3) per participant during the saline condition, 7 ICs (SD: 3) during the 0.5-μg dose, and 9 ICs (SD: 3) during the 2-μg dose. The number of removed ICs did not differ significantly between the groups during the saline condition (W=249, P=0.43, log(BF_10_)=-0.94; GAD: mean: 6, SD: 3; HC: mean: 5, SD: 2), or during the 2-μg dose (W=232, P=0.25, log(BF_10_)=-0.79; GAD: mean: 10, SD: 3; HC: mean: 9, SD: 3). During the 0.5-μg dose, a higher number of ICs were identified and removed in the GAD group than in the HC group (W=186, P=0.04, log(BF_10_)=0.28; GAD: mean: 8, SD: 4; HC: mean: 6, SD: 2).

### Supplementary Discussion

Theoretical insights from computational neuroscience suggest that interoception might be implemented differently than a neural process simply integrating internal state of the body via ascending afferent (i.e., “bottom-up”) pathways [19-21]. Unlike traditional hierarchical feedforward models of perception that involve a mostly linear filtering and translation of sensory signals to arrive at higher-order perception, a new argument has emerged that explains perception as arising from predictive processing [22]. In this perspective, neurons transmitting predictions about interoceptive states communicate with neurons detecting deviations from those predictions to develop an explanation for the perceptual information received (called a ‘generative model’) [19-21]. In the interoceptive context, a generative model is an internal model that specifies how interoceptive signals are generated from hidden states by incorporating prior (“top-down”) knowledge about the expected structure of the body (‘interoceptive priors’). Interoception (or aspects of it) may thus actually correspond to inferring the causes of internal signals through processes of inference, learning, and prediction according to certain forms of probability theory (specifically, Bayes’ theorem). According to this view, if the actual state of the body differs from the predicted state, this results in a prediction error (PE). When PEs are small or non-existent, the system can be said to be in homeostasis (i.e. balanced). By contrast, when PEs are large, incoming interoceptive information is decoupled from the brain’s interoceptive predictions, which may lead to dysfunction because of an inability to appropriately process salient environmental stimuli [23]. Computational theories of interoceptive psychopathology suggest that generalized anxiety disorder (GAD) results from abnormally strong expectations about situations eliciting bodily changes, which lead to mismatches between predictions and actual sensory inputs (i.e., larger interoceptive PE) [24]. Aberrant top-down mechanisms of interoceptive dysfunction in GAD may involve over-weighted expectations and failures to adjust them with actual physiological changes of the body.

Existing computational approaches to study cardiac interoception are associated with some methodological challenges. For instance, experimentally-induced perturbations of cardiac signal without influencing participant’s expectations require maskable manipulations that are challenging to implement in practice. The double-blinded intravenous administration of isoproterenol, a peripheral adrenergic agonist akin to adrenaline, provides an attractive experimental framework to modulate cardiovascular arousal while overcoming this limitation, thus contributing to facilitate investigation of top-down mechanisms underlying functional/dysfunctional processing of cardiac interoceptive signals [3, 25]. The present study was designed to achieve this goal. During the peak period of saline condition, participants may expect infusion-elicited changes in cardiac sensations, including increases in HR, which did not occur with saline infusion (Figure S8A). During this condition, the presence of over-weighted expectations about infusion-elicited bodily changes results in large PE. The HEP amplitude has been proposed to reflect the mismatch between expectations and actual cardiac inputs, based on the fact that top-down attention towards the heart, which increases PE by improving precision of cardiac input, modulates HEP amplitude [26], and the fact that PE are thought to be encoded by superficial pyramidal cells that are the major contributor to empirically observed neurophysiological responses [27, 28]. According to this interpretation, the larger HEP amplitude found in GAD group during the peak period of saline condition might reflect a larger mismatch between their expectations and their actual cardiovascular state, i.e., a larger PE, relative to healthy comparison (HC) group (Figure S12A). Even our interpretation is speculative, these findings echo recent computational theories of interoceptive psychopathology suggesting that abnormally strong expectations about situations eliciting bodily changes exert powerful influences on the emergence of dysfunctional avoidance behaviors in anxiety-related conditions [24, 29]. During low levels of adrenergic stimulation, specifically during the peak period of the 0.5-μg infusion, the isoproterenol-elicited increase in HR (Figure S2B) can be described as a stronger (increased signal-to-noise ratio) and more precise cardiac input, and the “distance” (from a Bayesian perspective) between the sensory input and the prior can be described as smaller in individuals with GAD who generally have abnormally strong expectations about situations eliciting bodily changes [24]. In other words, isoproterenol-induced cardiovascular changes could be associated with smaller PE in the GAD group, compared to HCs. Under this theoretical assumption, the distinct HEP amplitude patterns between the two groups during the 0.5-μg isoproterenol dose, which show small HEP amplitude in the GAD group and large amplitude in the HC group, may support the notion that HEP amplitude reflects interoceptive PE about cardiac signals (Figure S12B) [26, 27]. Our results also showed group differences in HEP amplitude during the early recovery period of the 0.5-μg dose of isoproterenol. According to a previous study characterizing attentional effects on HEP amplitude [26], the latency and electrode topography of the group difference in our study may be consistent with a pure attentional focus on the heart in the GAD group. From a physiological standpoint, the early recovery period of the 0.5-μg dose is characterized with the progressive decrease in HR (Figure S2B) that can be described as a weak (reduced signal-to-noise-ratio) cardiac input. In this context, the allocation of neural resources towards a cardiac sensory channel may be viewed as an attempt to increase the relative precision of incoming sensory information [30]. Our HEP results could be interpreted to suggest that, during the early recovery period of the 0.5-μg dose of isoproterenol, GAD participants (relative to HCs) directed a greater proportion of their neural attentional resource allocation to the heart to compensate for a less precise cardiac input as the cardiac activity neared baseline or resting levels. Such a top-down, cognitive strategy of attentional focus on the heart could be viewed as adaptative, particularly in light of empirical observations that interoceptive dysfunction in GAD can be characterized by a lower precision of ascending cardiac signals [31].Future studies may investigate the degree to which aberrant top-down processing of cardiac signals in GAD is implemented across response modalities (i.e., neural circuits, physiology, behavior, and self-report) using computational models of cardiac interoception [19]. For instance, Smith et al (2020) used heartbeat tapping task to provide behavioral evidence that individuals with symptoms of anxiety and comorbid depression/anxiety do not show hyperprecise interoceptive priors [31]. Extending computational model of cardiac interoception to the HEP measure during maskable manipulation of cardiovascular arousal may represent an important step to inform top-down neural underpinnings of interoceptive dysfunction in GAD [24]. Computational theories of interoceptive psychopathology in GAD may also be evaluated by testing the existence of a statistical relation between the HEP amplitude measured within the context of heartbeat tapping task and the parameters of computational model of cardiac interoception, as estimated from behavioral measures [31]. A statistically significant association between HEP amplitude and model parameters, specifically the PE, would be considered as direct evidence supporting the interpretation of the HEP amplitude as an index of PE about cardiovascular state [26, 27].

---

**Table S1.** Psychotropic medication status, diagnostic comorbidities, and race and ethnicity of general anxiety disorder (GAD) and healthy comparison (HC) participants.

|  | **GAD**<br>**(n=24)** | **HC**<br>**(n=24)** |
| --- | --- | --- |
|  | **N (%)** |  |
| **Psychotropic medication<sup>(a)</sup>** | 6 (25) | 0 |
| **Comorbid Diagnoses<sup>(b)</sup>** |  |  |
| Major depressive disorder (recurrent) | 8 (33) | 0 |
| Major depressive disorder (partial remission) | 6 (25) | 0 |
| Social anxiety disorder | 8 (33) | 0 |
| Posttraumatic stress disorder | 2 (8) | 0 |
| **Race and Ethnicity<sup>(c)</sup>** |  |  |
| Black or African American | 1 (4) | 0 |
| American Indian or Alaskan Native | 1 (4) | 1 (4) |
| Asian | 3 (13) | 4 (17) |
| Hispanic or Latino | 3 (13) | 2 (8) |
| White | 19 (79) | 18 (75) |
| Other | 0 | 0<sup>(d)</sup> |

N: number of participants; %: proportion of participants
<sup>(a)</sup> 4 participants reported selective serotonin reuptake inhibitor use (sertraline, paroxetine, fluoxetine), 1 reported selective norepinephrine reuptake inhibitor use (escitalopram), 1 reported taking both classes of reuptake inhibitor, 1 reported occasional as needed use of a short-acting beta-blocker (propranolol) but abstained for at least one day prior to scanning and 1 reported medicinal tetrahydrocannabinol (THC) use but tested negative on the day of scanning. No participants were taking scheduled benzodiazepines or barbiturates.
<sup>(b)</sup> For brevity, only comorbid diagnoses with frequency > 5% are listed.
<sup>(c)</sup> Racial and ethnic categories defined as per the most recent revisions of the federal Office of Management and Budget Directive 15.
<sup>(d)</sup> One healthy comparator refused to report race and ethnicity.

**Table S2.** A descriptive and approximate classification scheme for the interpretation of the log scale of Bayes factor (Log (BF_10_), adapted from Jeffreys, 1961 [32].

| | *Log* ($BF_{10}$) | Interpretation | Symbol |
| --- | --- | --- | --- |
| Evidence favoring $H_1$ ↑ | > 2 | extreme evidence for $H_1$ | $H_1^{****}$ |
| | 1.48 to 2 | very strong evidence for $H_1$ | $H_1^{***}$ |
| | 1 to 1.48 | strong evidence for $H_1$ | $H_1^{**}$ |
| | 0.48 to 1 | moderate evidence for $H_1$ | $H_1^*$ |
| | 0 to 0.48 | anecdotal evidence for $H_1$ | ns |
|  | 0 | no evidence | ns |
| Evidence favoring $H_0$ ↓ | -0.48 to 0 | anecdotal evidence for $H_0$ | ns |
| | -1 to -0.48 | moderate evidence for $H_0$ | $H_0^*$ |
| | -1.48 to -1 | strong evidence for $H_0$ | $H_0^{**}$ |
| | -2 to -1.48 | very strong evidence for $H_0$ | $H_0^{***}$ |
| | < -2 | extreme evidence for $H_0$ | $H_0^{****}$ |

*log*( $BF_{10}$ ): log scale of Bayes factor $BF_{10}$ ; $H_1$ : alternative hypothesis; ns: non-significant; $H_0$ : null hypothesis

**Table S3.**
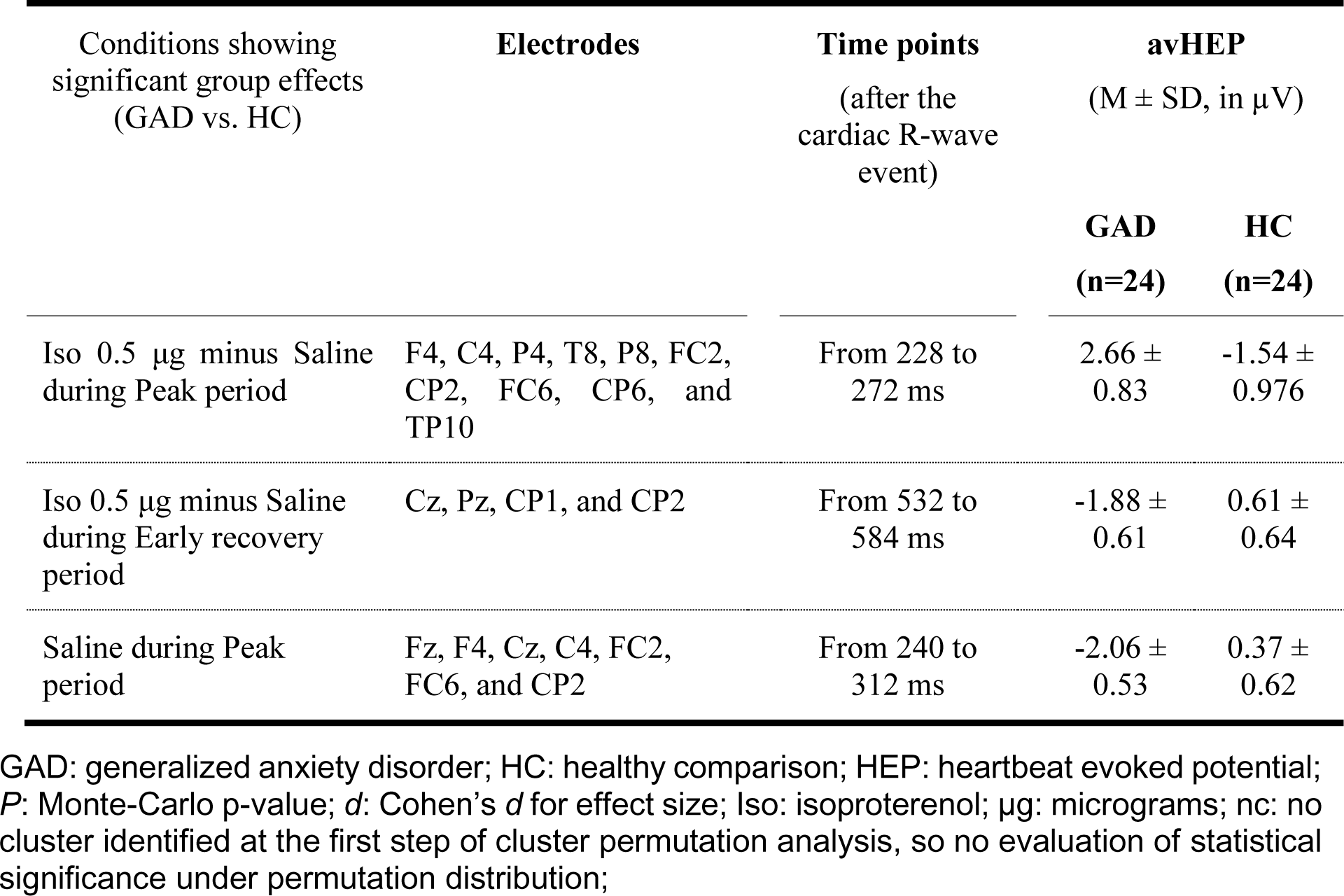
Statistics from cluster permutation analysis informing on how HEP amplitudes differed between the GAD and HC groups, based on various comparisons made both within and between conditions.

**Figure S1.**
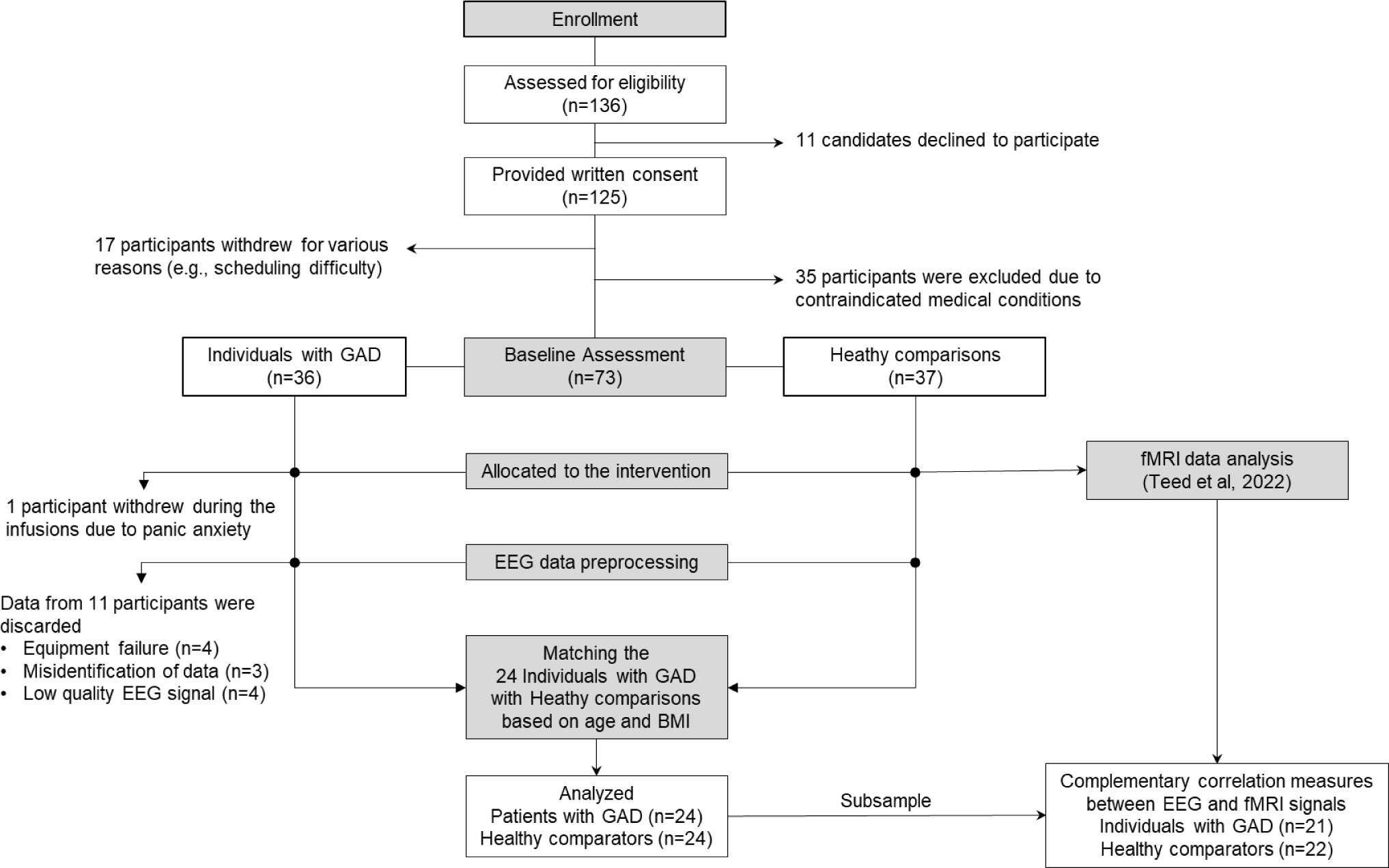
CONSORT flow diagram of the study. GAD: generalized anxiety disorder; EEG: electroencephalogram; BMI: body mass index; fMRI: functional magnetic resonance imaging

**Figure S2.**
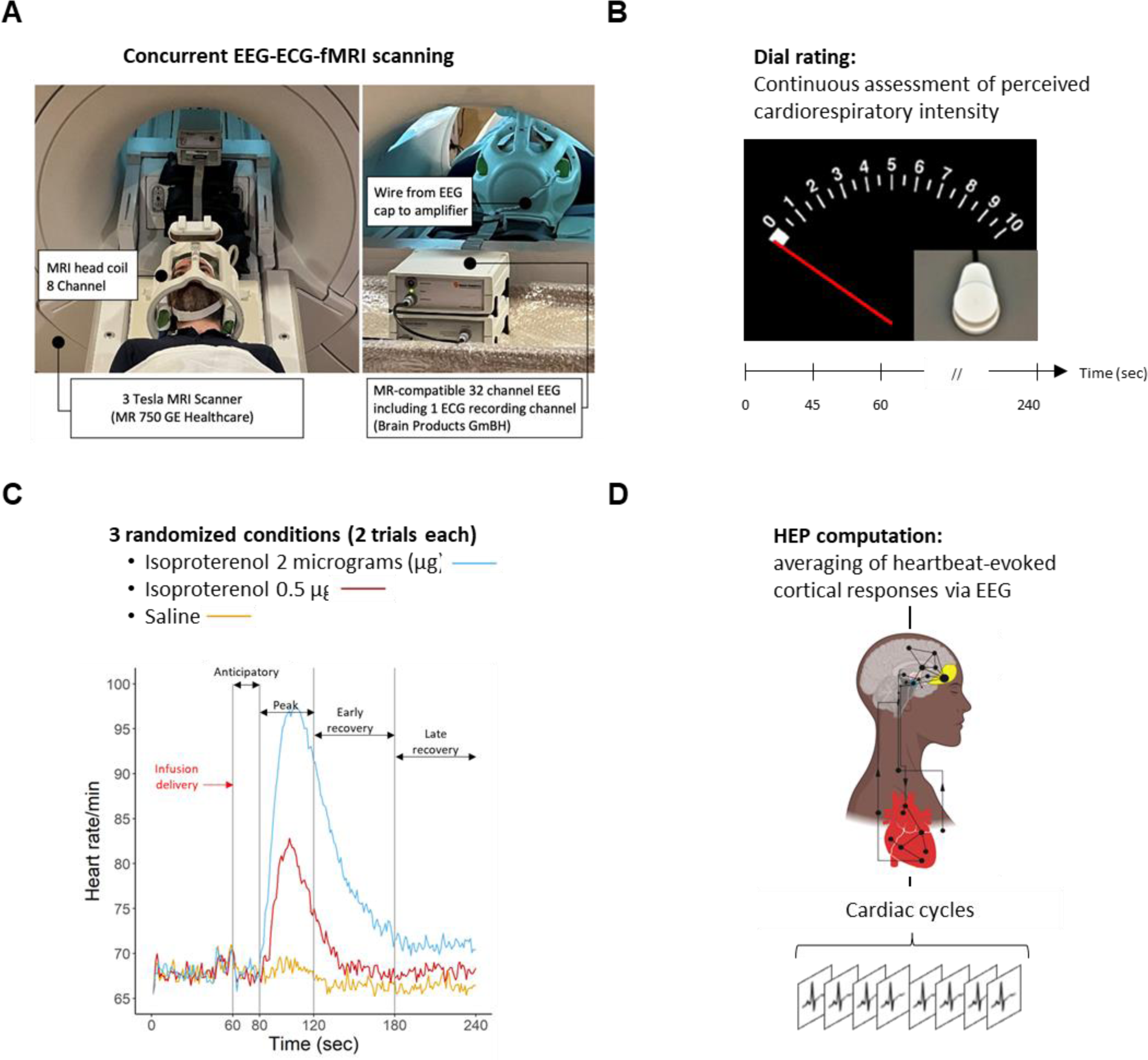
**(A)** Experimental setup for simultaneous recording of neural (electroencephalogram, EEG), cardiovascular (electrocardiogram, ECG), and subjective responses to peripheral adrenergic stimulation with isoproterenol during functional magnetic resonance imaging (fMRI) scanning. Participants wore a 32-Channel MRI compatible EEG cap while MRI signals were collected from an 8-channel head coil. **(B)** During each scan, participants continuously rated changes in the perceived intensity of cardiorespiratory sensations by rotating an MRI-compatible dial from 0 (none or normal) to 10 (most ever). **(C)** Average heart rate responses to isoproterenol and saline across our sample of 24 individuals with Generalized Anxiety Disorder (GAD) and 24 Healthy Comparators (HC). Bolus infusions were delivered at 60 seconds after the start of each infusion scan. For illustrative purpose, key infusion periods (i.e., anticipatory, peak, early and late recovery) are superimposed on the graph. **(D)** The Heartbeat Evoked Potential (HEP) was computed by averaging the EEG signal, which was synchronized on the cardiac R-wave event of the ECG, across cardiac cycles. Μg = micrograms, sec = seconds.

**Figure S3.**
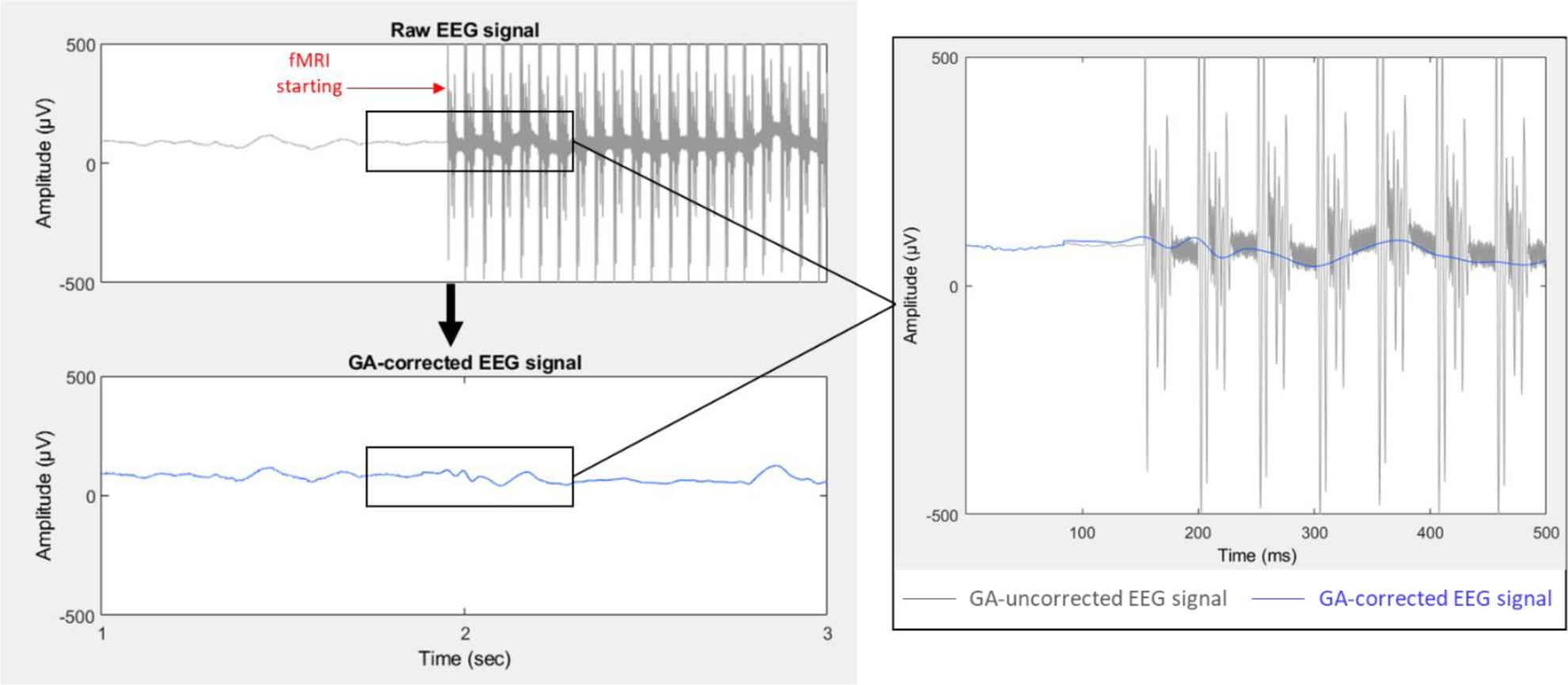
Procedure for gradient artifact (GA) removal from the electroencephalogram (EEG) signal collected during fMRI scanning. The Optimal Basis Set (OBS) method was implemented to correct for the GA using the fMRIb plug-in (version 2.00) for EEGLAB (Niazy et al, 2005). The right subplot shows a representative 500 millisecond (ms) example of an EEG signal from the O2 electrode before (dark grey) and after (blue) correction of seven GA artifact sequences using OBS. μV – microvolts, sec – seconds, ms – milliseconds

**Figure S4.**
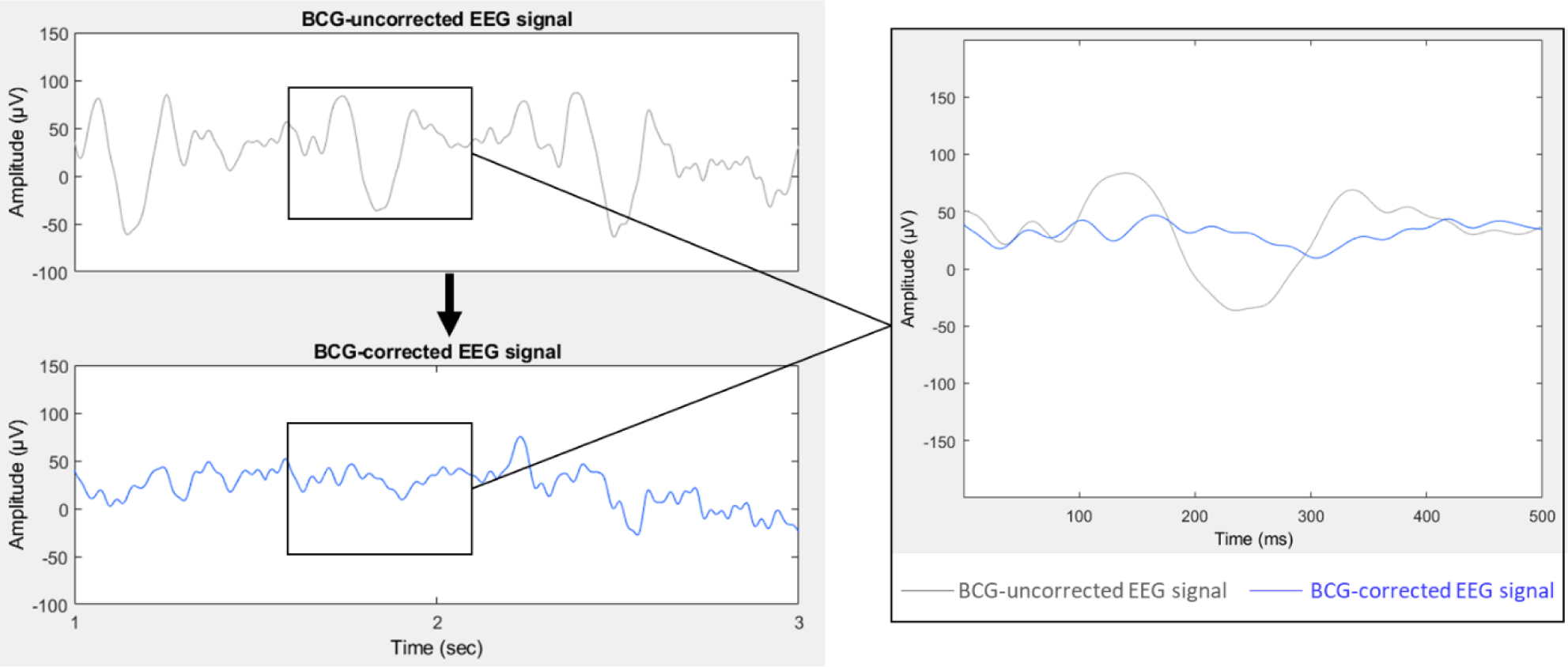
Procedure for removing the ballistocardiogram artifact (BCG) from the electroencephalogram (EEG) signal collected during fMRI scanning. The Optimal Basis Set (OBS) method was implemented to correct for the BCG artifact using the fMRIb plug-in (version 2.00) for EEGLAB (Niazy et al, 2005). The right subplot shows a representative 500 millisecond (ms) example of an EEG signal from the CP6 electrode before (dark grey) and after (blue) a single BCG artifact correction using OBS. μV – microvolts, sec – seconds, ms – milliseconds

**Figure S5.**
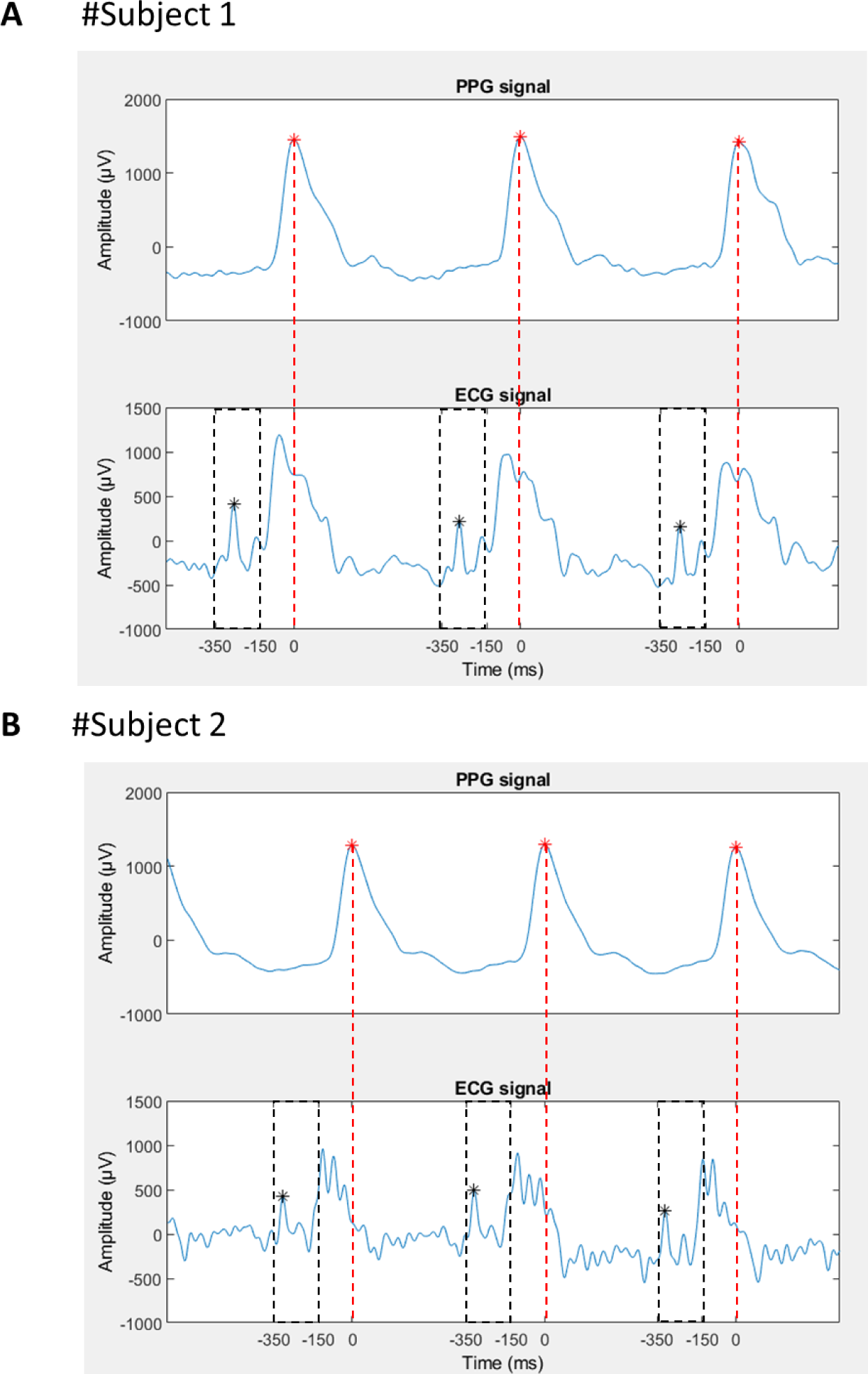
Illustration of the successful identification of cardiac R-wave events on the ECG signal collected during MRI scanning via a semi-automated approach using additional information from the photoplethysmogram (PPG) signal collected from a nondominant finger. ECG R peaks were automatically detected within a -350 to -150 ms time window (dashed black rectangle) before the PPG waveform peak, based on previously published findings showing a delay of approximately 250 ms between the ECG R peak (black asterisk in lower plots of panels A and B) and the finger PPG waveform peak (red asterisk in upper plots) [7, 8]. ECG data were subsequently visually inspected to assure that the cardiac R wave events were correctly identified. Panels A and B illustrate the interindividual variability of ECG signals following gradient artifact correction. While Subject 2 shows more variability in the signal than Subject 1, in both cases the R-peak could be reliably identified. μV – microvolts, ms – milliseconds

**Figure S6.**
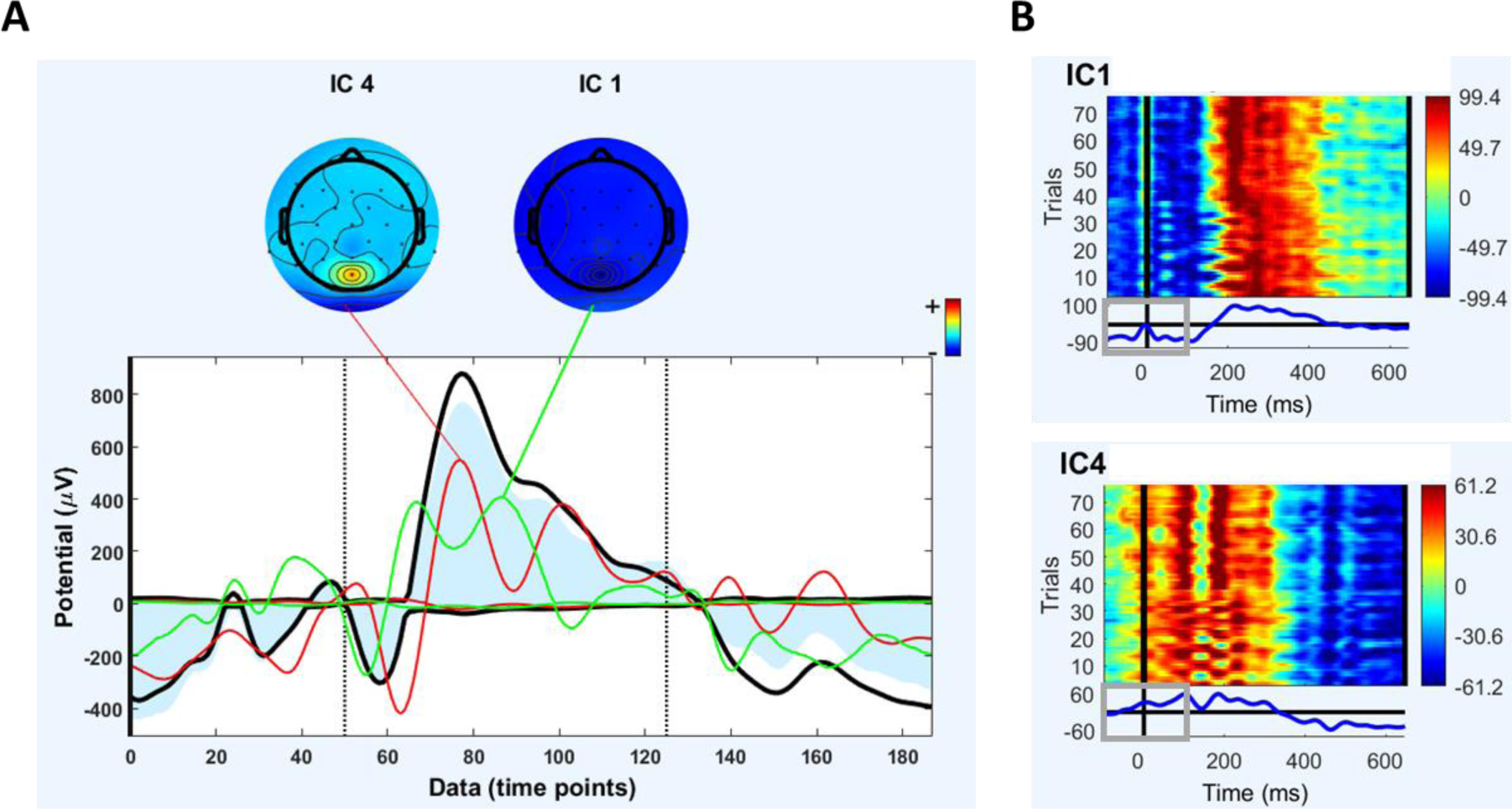
**(A)** A typical example of two independent components that were identified as related to ballistocardiogram artifact residuals, based on the amount of variance they contributed to the evoked pulse artifact, using the EEGLAB function eeg_pvaf() [9, 10]. The two independent components accounted for >93% of the variance of the evoked pulse artifact in the time range. Shown are the envelope of the sensor data (in black), which reflects the minima and maxima across all channels, and the envelope of the joint back-projection of the two independent components (blue shaded area). For each independent component, the map (i.e., inverse weights) and the envelope of the back-projection (colored traces) are shown. **(B)** Trial-by-trial view (one trial corresponds to one heart cycle) of the time course for two independent components that were identified as related to the cardiac field artifact (CFA), because their maximum amplitude was higher than 5 μV within the time window [-100 100 ms] from the cardiac R-wave event (time zero), and their timing was similar across trials (heart cycles). μV – microvolts, ms – milliseconds

**Figure S7.**
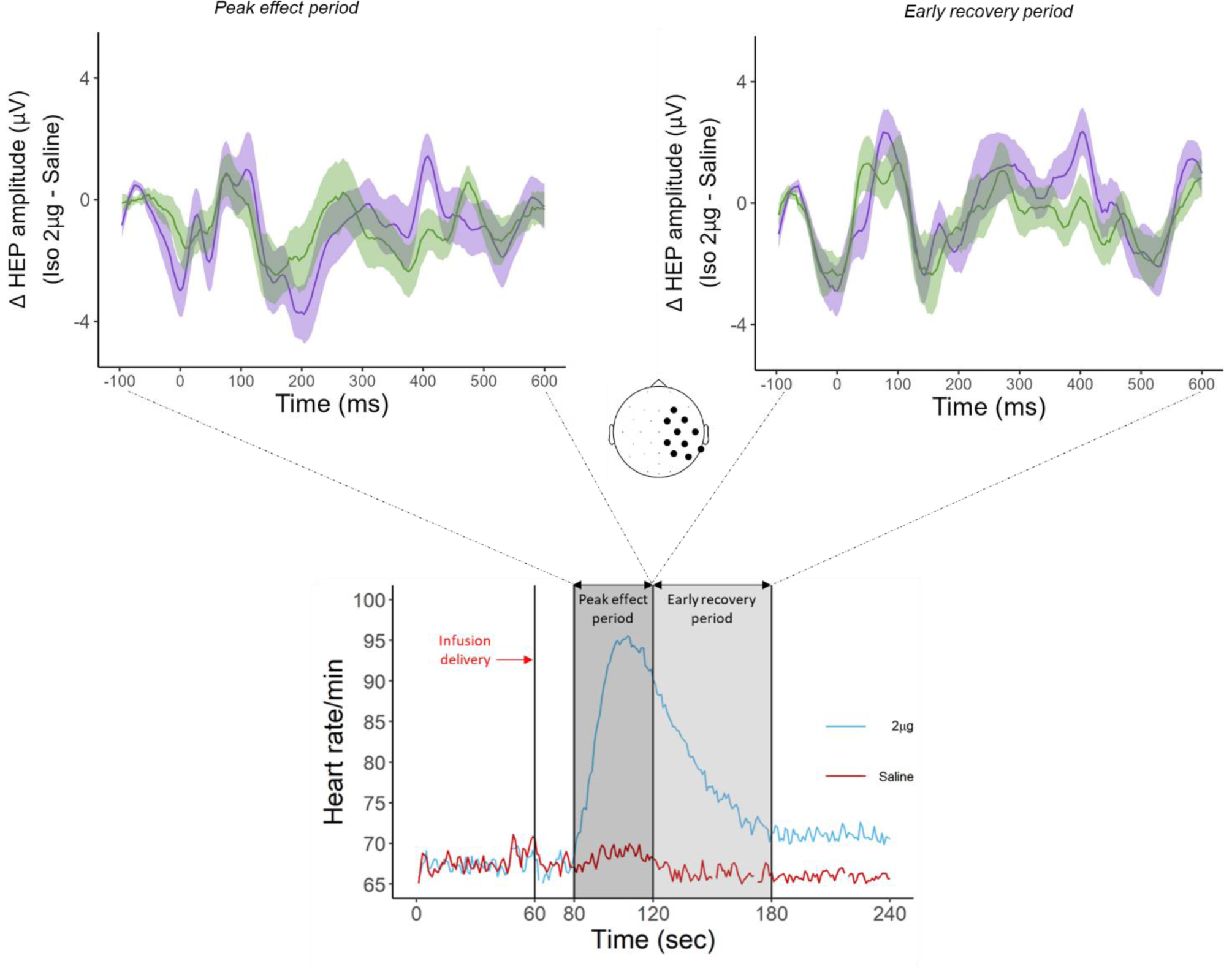
Changes in HEP amplitude between the 2-μg and saline infusions during the peak and the early recovery periods. There were no significant group differences observed for either period*. Note: The HEP was computed within the cluster of right frontocentral and parietal electrodes identified as showing group differences between the 0.5-μg and saline infusions during the peak period (i.e., Figure 1A).* GAD – generalized anxiety disorder, HC – healthy comparison, Δ – delta, μg – micrograms, μV – microvolts, ms – milliseconds, sec – seconds

**Figure S8.**
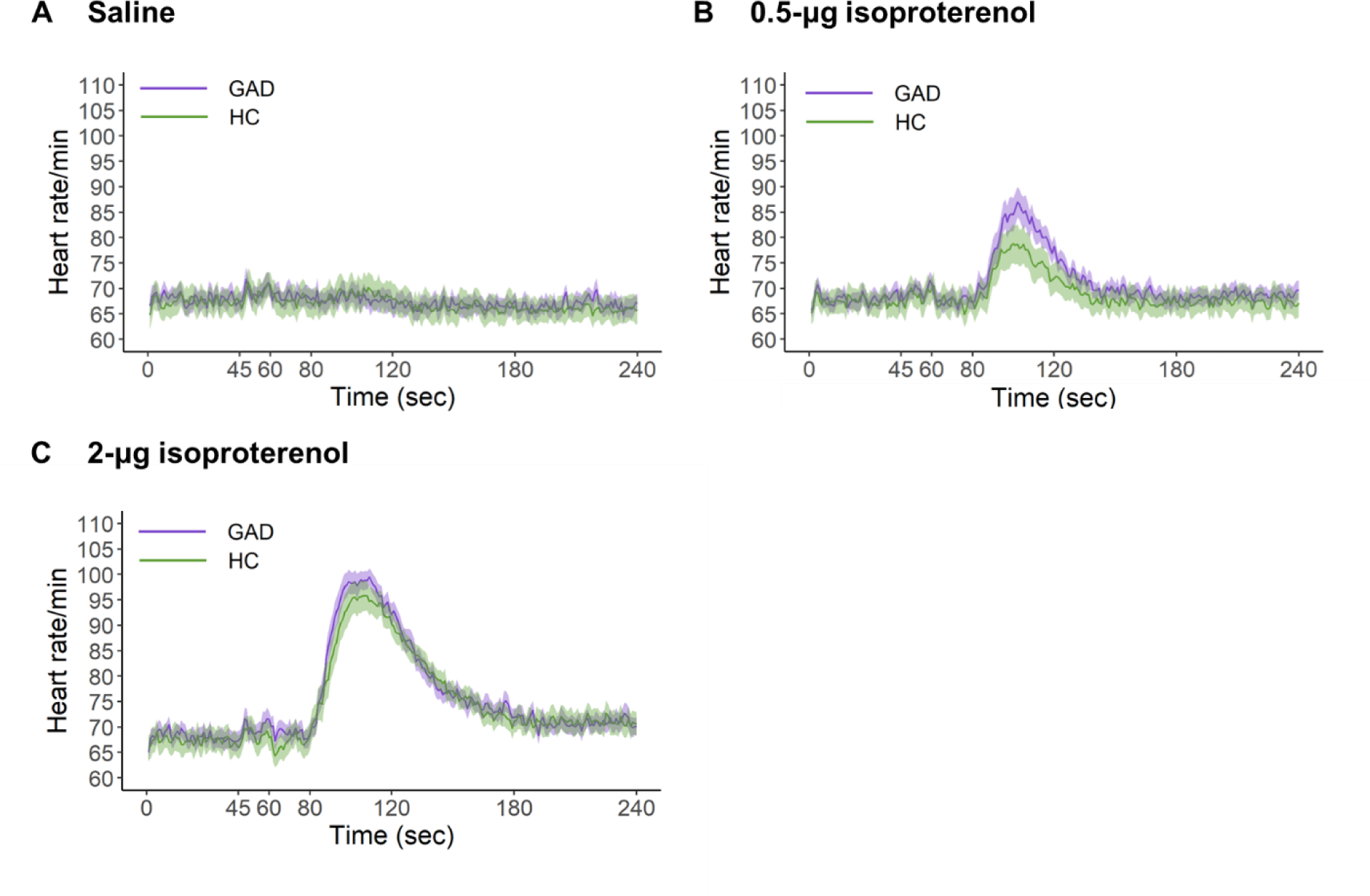
Mean heart rate during **(A)** the saline infusion, **(B)** the 0.5-μg infusion of isoproterenol, and **(C)** the 2-μg infusion of isoproterenol. No significant group difference was observed when analyzing heart rate responses during the three infusions. GAD – generalized anxiety disorder, HC – healthy comparison, μg – micrograms, sec – seconds.

**Figure S9.**
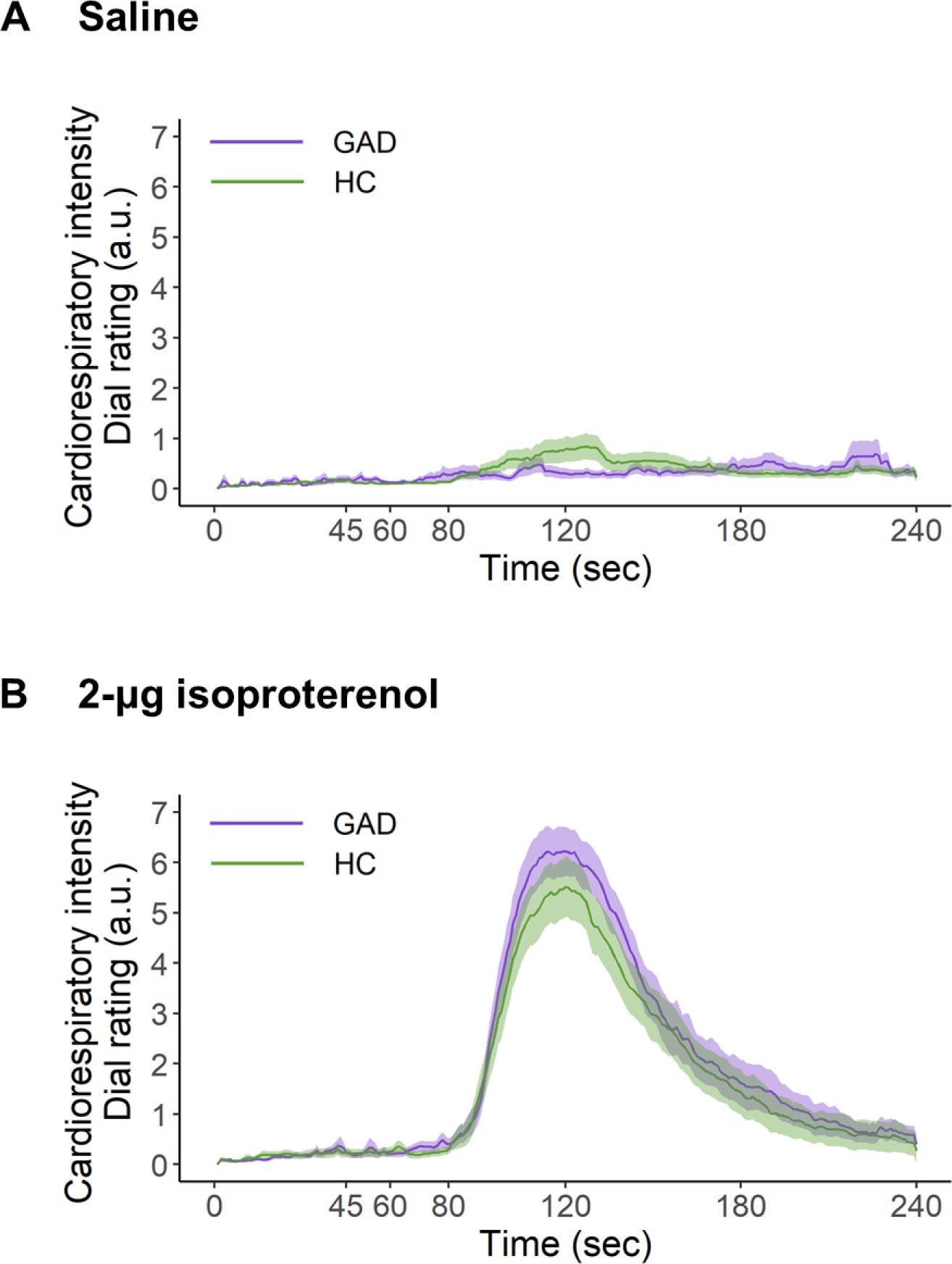
Mean dial ratings of cardiorespiratory sensation intensity during **(A)** the saline infusion and **(B)** the 2-μg infusion of isoproterenol. No significant group effect was found when analyzing dial responses during the saline and the 2.0-μg infusions. GAD – generalized anxiety disorder, HC – healthy comparison, μg – micrograms, a.u. – arbitrary unit, sec – seconds.

**Figure S10.**
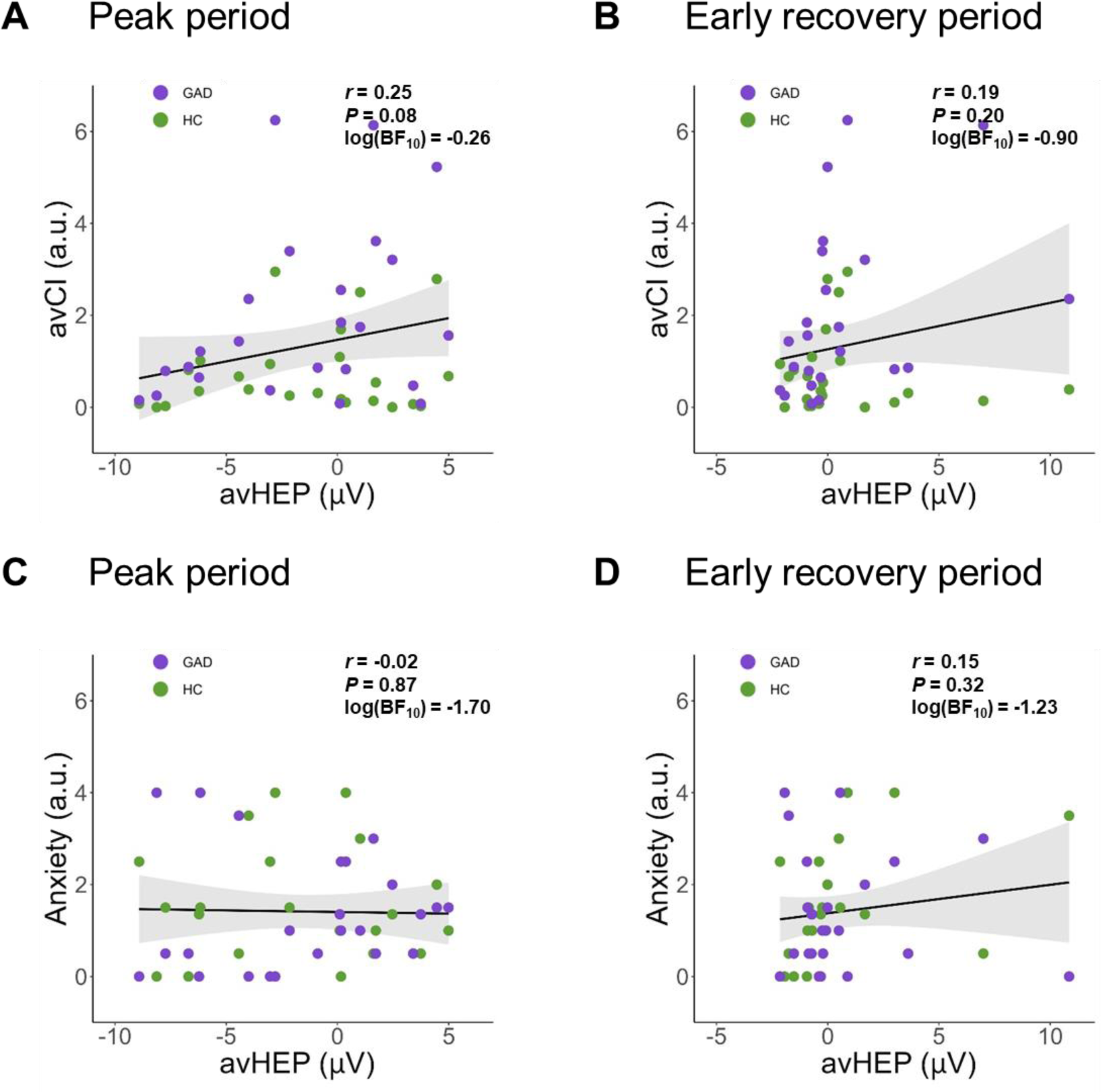
Statistical independence between the average HEP amplitude (avHEP), computed as the average HEP signal calculated across electrodes × time points that were included in the HEP cluster of the peak period and the early recovery period (see Figure 1 in the main text) and the average self-reported cardiorespiratory intensity (avCI) **(A-B)**, as well as the self-reported anxiety **(C-D)**. GAD – generalized anxiety disorder, HC – healthy comparison, μV – microvolts, a.u. – arbitrary unit

**Figure S11.**
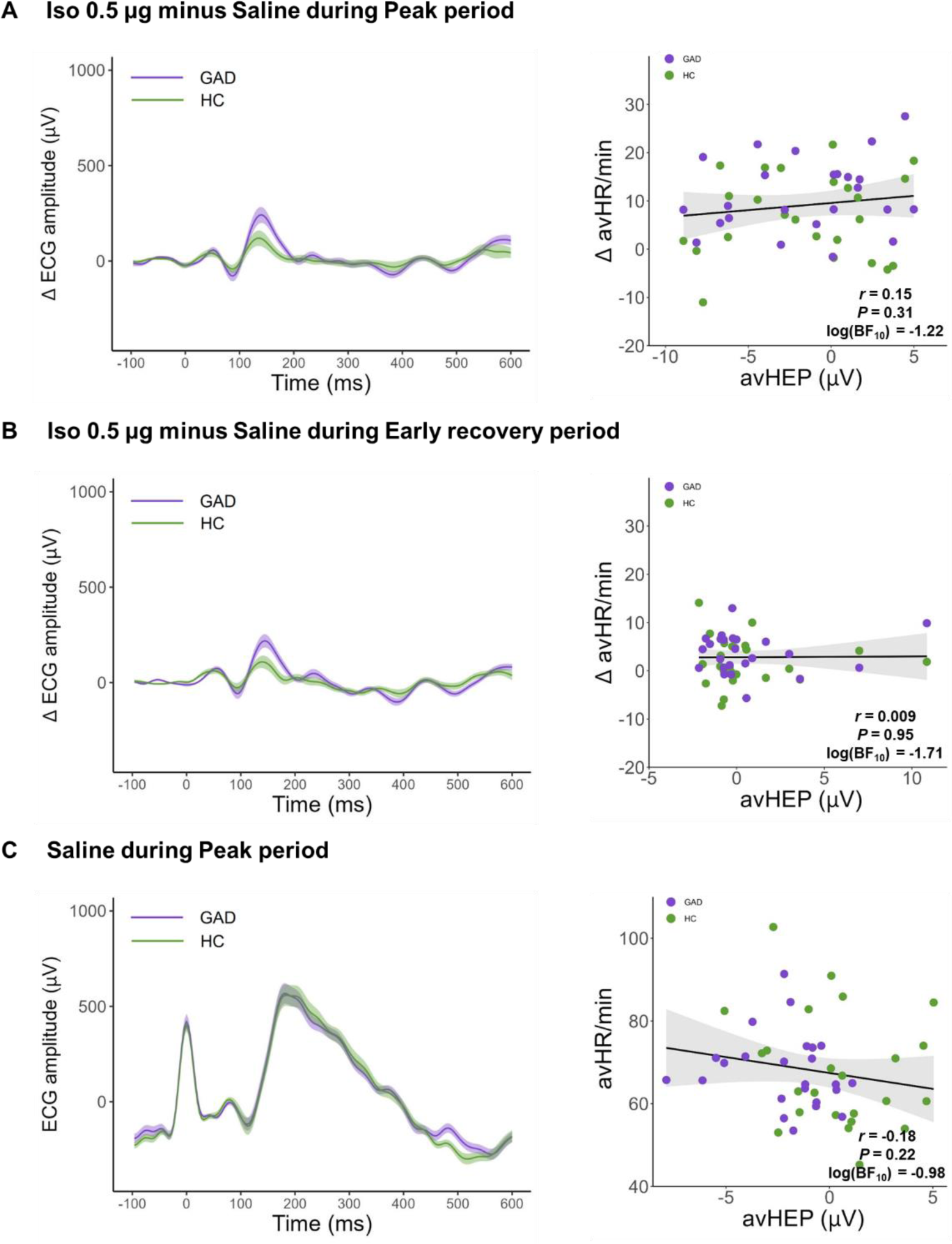
Changes in cardiac signal from the saline to the 0.5-μg infusions, during **(A)** the peak period and **(B)** the early recovery period. **(C)** Cardiac signal during the peak period of the saline infusion. **(A-C, left panels)** No significant group difference was found when analyzing the change in ECG amplitude between the saline and the 0.5-μg infusions, and the ECG amplitude in the saline infusion. **(A-C, right panels)** The average HR (avHR) computed across all time points of the period of interest was uncorrelated with the average HEP amplitude (avHEP), as calculated across electrodes × time points that were included in the corresponding HEP cluster. GAD – generalized anxiety disorder, HC – healthy comparison, Δ – delta, μg – micrograms, μV - microvolts, ms – milliseconds, sec – seconds, min - minutes.

**Figure S12.**
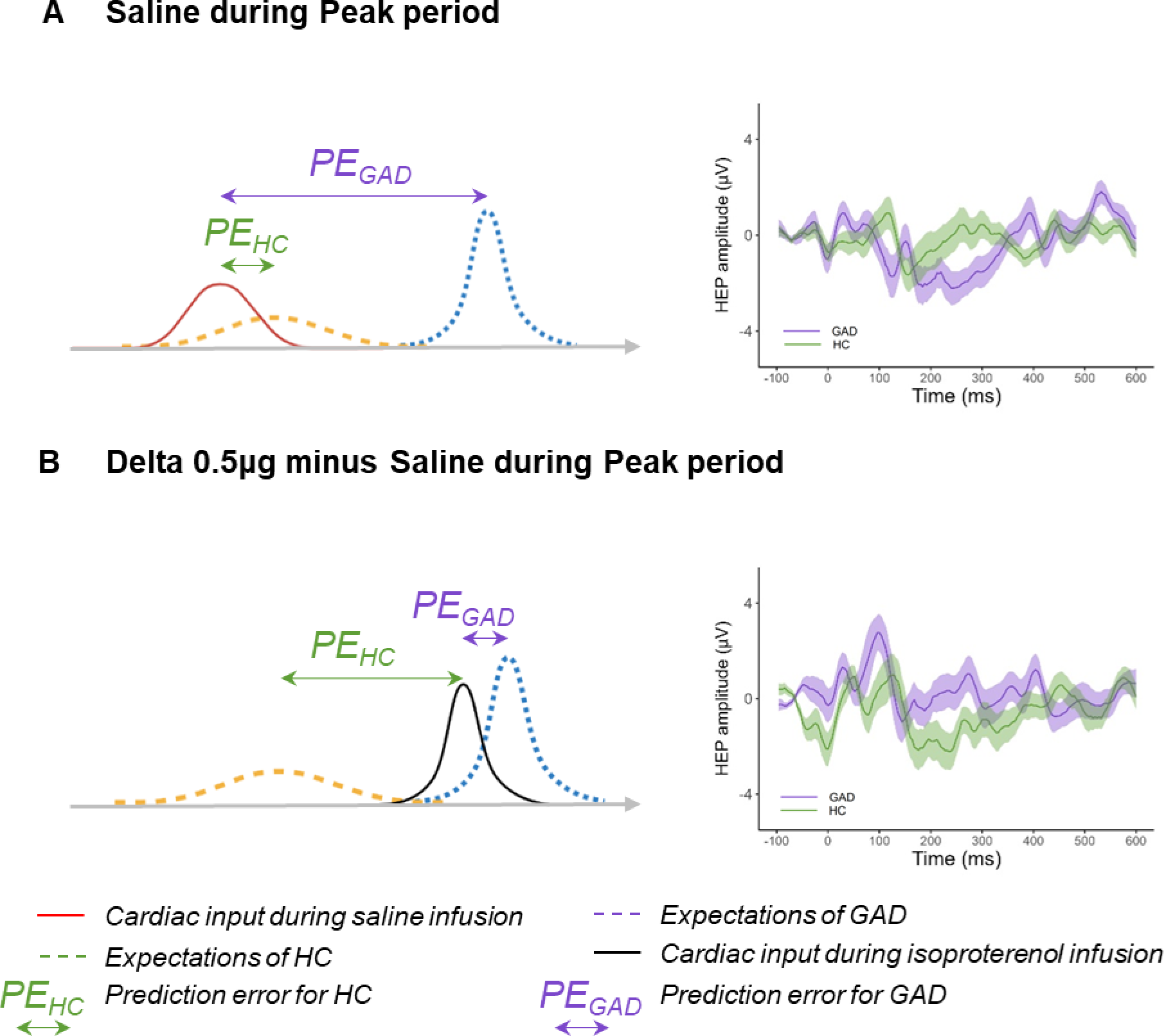
Graphical summary of the discussion regarding putative relations between HEP findings of the present study and computational models of interoception and psychopathology [19-21, 24]. **(A)** During the saline condition, participants may expect infusion-elicited changes in cardiac sensations (e.g., increase in heart rate (HR)) that did not occur with saline infusion, thus generating a prediction error (PE) signal. According to the interpretation of the HEP amplitude as an index of PE [26, 27], the larger HEP amplitude found in individuals with generalized anxiety disorder (in purple; GAD) could reflect a larger PE, namely a larger mismatch between their expectations and their actual cardiovascular state to relative to healthy comparators (in green; HC). **(B)** During low levels of adrenergic stimulation, one can argue that isoproterenol-elicited (Iso) increase in HR make cardiac input precise (high signal-to-noise ratio), and its “distance” (from a Bayesian perspective) from the prior can be described as smaller in individuals with GAD because they generally have abnormally strong expectations about situations eliciting bodily changes [24]. Under this theoretical assumption, the distinct HEP amplitude patterns between the two groups during the 0.5-μg isoproterenol dose, which show small HEP amplitude in the GAD group and large amplitude in the HC group, may support the notion that HEP amplitude reflects PE about cardiac signals [26, 27].

